# Effectiveness of pneumococcal vaccination campaigns in humanitarian settings: a modelling study

**DOI:** 10.1101/2025.05.16.25327803

**Authors:** Kevin van Zandvoort, Mohamed O Bobe, Abdirahman I Hassan, Rachael Cummings, Abdihamid Warsame, Casey L Pell, Mohamed Ismail Abdi, Saed Ibrahim, Catherine R McGowan, E Kim Mulholland, Catherine Satzke, Rosalind M Eggo, Mohamed Abdi Hergeye, Francesco Checchi, Stefan Flasche

**Affiliations:** Department for Infectious Disease Epidemiology, London School of Hygiene and Tropical Medicine, United Kingdom; Health and Nutrition, Save the Children International, Somaliland; Republic of Somaliland Ministry of Health Development, Somaliland; Save the Children, United Kingdom; Infection, Immunity and Global Health, Murdoch Children’s Research Institute, The University of Melbourne Department of Paediatrics at the Royal Children’s Hospital, Australia; Department of Microbiology and Immunology, The University of Melbourne at the Peter Doherty Institute for Infection and Immunity, Australia; Centre for Global Health, Charité - Universitätsmedizin Berlin, Germany

## Abstract

**Background:** A large and increasing number of people are forcibly displaced worldwide because of war, food insecurity, and other crises. *Streptococcus pneumoniae* (the pneumococcus) likely causes a substantial health burden in crisis-affected populations, but pneumococcal vaccines (PCVs) are rarely used in humanitarian responses. We evaluated the potential impact of logistically feasible PCV campaigns in such settings.

**Methods:** We conducted a pneumococcal carriage, malnutrition and social contact survey in a camp for displaced people near Hargeisa, Somaliland, to parameterise a transmission model accounting for migration. We projected the effect of PCV mass-vaccination campaigns using one or two doses and vaccinating children <1, 2, 5, 10 or 15 years (y) of age in the Somaliland camp and three different crisis settings.

**Findings:** A single-dose PCV campaign with high vaccine coverage in children <5y or older can partially control vaccine-serotypes for up to three years, preventing 27% (95%CrI 20-34) of severe pneumococcal disease when vaccinating <5y olds and 38% (95%CrI 29-46) when vaccinating <15y olds. Expanded age eligibility is needed for comparable protection in settings with increased migration or more interaction with unvaccinated host populations. A campaign vaccinating <5y olds is the most efficient use of PCV with 108 (95%CrI 77-162) vaccines needed to prevent one case of severe pneumococcal disease. Implementing such campaigns in displaced populations worldwide would require about 40 million doses in the next five years.

**Interpretation:** Single-dose PCV mass-vaccination campaigns offer crisis-affected populations an effective, pragmatic immunisation strategy.

**Funding:** This study was funded by Elrha’s Research for Health in Humanitarian Crises (R2HC) Programme.

**Research in context:** *Evidence before this study:* Acute respiratory infections are a leading cause of morbidity and mortality in crisis-affected populations, but pneumococcal conjugate vaccines are rarely used in humanitarian responses. Routine vaccination is often not feasible in these settings due to a lack of access or security, and there is little guidance on alternative delivery options for these vaccines. A literature search on Apr 10, 2025, using the terms (“pneumococcal conjugate vaccine*” OR “PCV”) AND (“humanitarian” AND (“cris*” OR “emergenc*”)) returned 9 results on Embase and 8 results on PubMed. PCV mass vaccination strategies have previously been shown to be cost-effective in crisis settings, but no study has directly compared the effect and efficiency of different age eligibility and dosing regimens in crisis-affected populations.

*Added value of this study:* We collected key primary data that would allow us to parameterise a model to project the effect of different PCV campaigns on the transmission of vaccine-type pneumococci. To our knowledge, this is the first analysis that has assessed the combined direct and indirect effect of PCV campaigns in crisis-settings. We show the importance of achieving high levels of indirect protection for a substantial and sustained impact, which can be realized by vaccinating the main transmitters in addition to the children at highest risk of severe disease. We show that this can likely be achieved using a single-dose strategy, which greatly improves the feasibility of a campaign.

*Implications of all the available evidence.:* Single-dose mass-vaccination PCV campaigns that achieve high coverage in children up to 14 years (y) of age can provide substantial health benefits in crisis-affected populations. Vaccinating <5y olds may be most efficient. Temporarily disrupting transmission of vaccine-type pneumococci is crucial to protect unvaccinated children born or in-migrated after the campaign.

## Introduction

In 2024, over 120 million people, including more than 48 million children, were forcibly displaced worldwide due to war or conflict, food insecurity, natural disasters, and other crises.^1^ Displaced populations often live in overcrowded settings where infectious disease outbreaks are common, with poor access to healthcare and increased levels of morbidity and mortality. *Streptococcus pneumoniae* (the pneumococcus) likely causes a sizeable proportion of this burden,^2,3^ but pneumococcal conjugate vaccines (PCVs) are rarely used during humanitarian responses.^4^

The WHO recommends to use PCVs in crisis settings in children <1y old, and to consider their use in children <5y old.^5,6^ These ages align with routine vaccination schedules and campaigns used during PCV introduction into the Expanded Programme on Immunization. However, crisis settings may require alternative vaccination strategies, as limited access to services, disruption of cold or supply chains, lack of sufficient trained personnel and/or reduced safety of health care workers can make routine vaccination challenging to implement.^4^

Mass vaccination campaigns (MVCs) are commonly used in humanitarian responses to rapidly provide direct and indirect protection against pathogens causing diseases such as measles, polio, and cholera.^7^ MVCs often give a single-dose of vaccine to many age groups as a pragmatic approach to maximize vaccination coverage^8^. This not only immunizes those most at risk of severe disease, but could also interrupt transmission by leveraging indirect protection (herd immunity). Sufficient levels of herd immunity could i) mitigate suboptimal direct protection induced by an incomplete vaccination schedule in infants, and ii) extend protection to unvaccinated cohorts of newborns or newly migrated individuals who entered the population after an MVC. This is especially relevant in crisis-affected settings with high birth rates and migration.^4^

To assess the impact of PCV vaccination strategies that may feasibly be used in crisis settings, we previously collected pneumococcal carriage^9^ and risk factor data^10^ in a camp (Digaale) for internally displaced people (IDP) in Somaliland to inform a pneumococcal transmission model. We here report our modelled findings of the effectiveness and efficiency of potential PCV campaigns in Digaale and three additional exemplar crisis settings.

## Methods

### Transmission Model

We developed a compartmental model to simulate pneumococcal transmission within and between a displaced and host population. We grouped pneumococcal serotypes as vaccine-type (VT) or non-vaccine-type (NVT), depending on their inclusion in the Pneumosil vaccine (serotypes 1, 5, 6A, 6B, 7F, 9V, 14, 19A, 19F, and 23F), a low-cost PCV targeting serotypes commonly circulating in low- and middle-income countries. Populations were stratified by age, malnutrition status and vaccination status. In our model, susceptible individuals (S) can become infected with VT or NVT following effective contact with a VT or NVT carrier (Figure 1). Individuals carrying VT or NVT can acquire a superinfection (heterologous carriage; B) through additional infection with the other serotype group. Acquisition of superinfection occurs at a lower rate than acquisition of a first infection for susceptible people, as serotypes compete. More details including model equations are provided in supplemental material section A.

**Figure 1.**
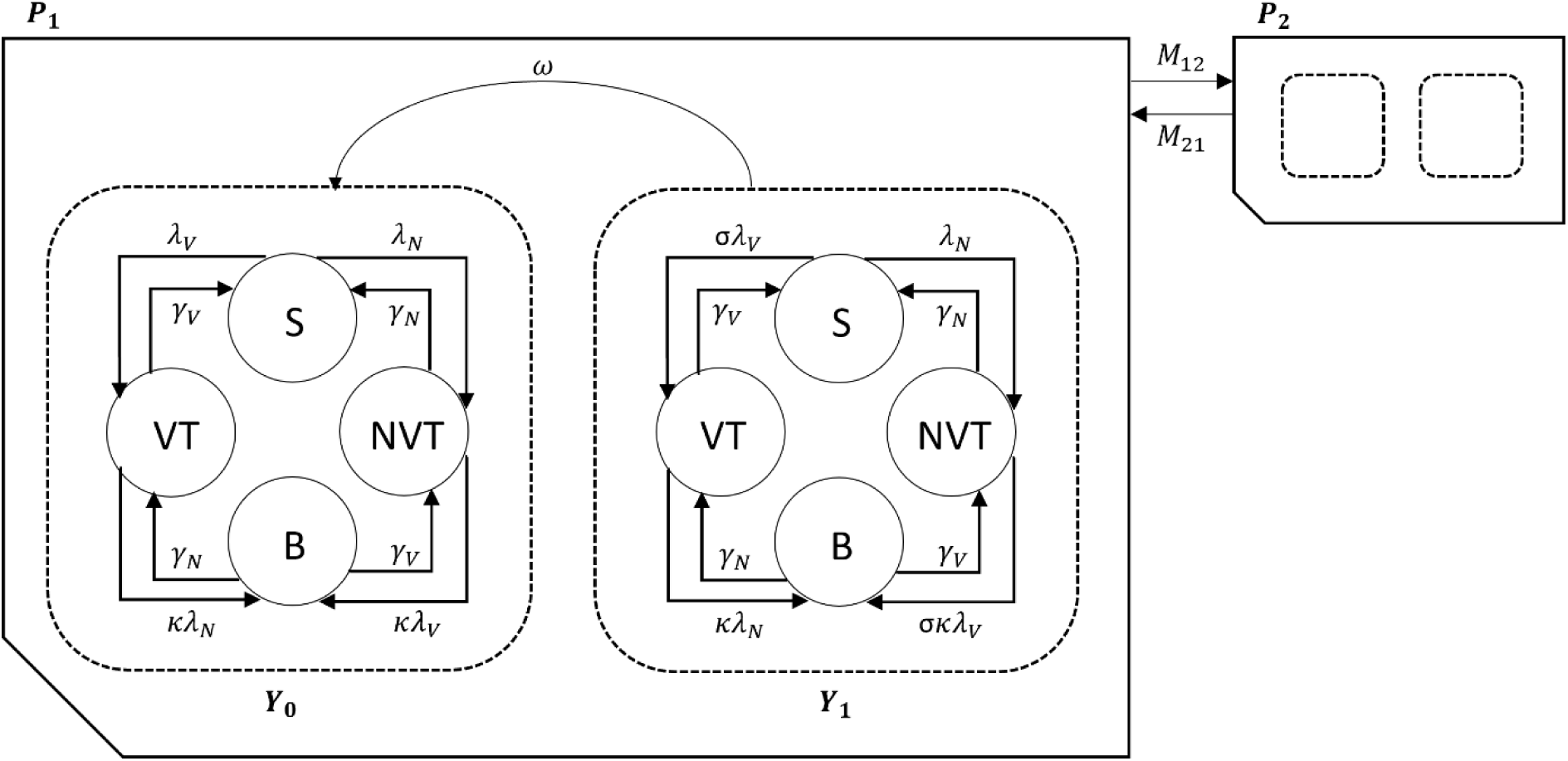
Structure of the compartmental pneumococcal transmission model. Pneumococcal serotypes are grouped as vaccine-type (VT), or non-vaccine-type (NVT). At any time *t*, susceptible individuals (S) acquire serotypes in either group at rates *λ*_*v*_(*t*) and *λ*_*N*_(*t*), and become infectious carriers themselves. Carriers of serotypes in one group remain partially susceptible to infection with serotypes in the other group, and may develop a superinfection (B) at rates *κλ*_*v*_(*t*) and *κλ*_*N*_(*t*), where *κ* models the reduced acquisition rate due to competition between serotypes. Carriers are assumed to clear their infection at rates *γ*_*v*_ for VTs and *γ*_*N*_ for NVTs. Individuals may be with (*Y*_1_) or without vaccine derived protection (*Y*_0_). Those with vaccine protection acquire VTs at a reduced rate, *σ*, and lose their vaccine protection at rate *ω*. Different populations *P*_1_ and *P*_2_ represent a displaced and host population. Individuals of population *p* migrate to population *q* at rate *M_qp_*. Multiple age groups, ageing, malnourished strata, and transmission between these groups are not shown.

We implemented mixing between age groups as an age-dependent daily contact rate (Table 1), informed by setting-specific contact matrices from the Digaale IDP camp^10^ and other crisis-affected populations (described below). Contacts between the modelled populations are governed by a travel matrix, which incorporates a constant parameter for the proportion of extra-household contacts made with individuals in each population.

**Table 1.**
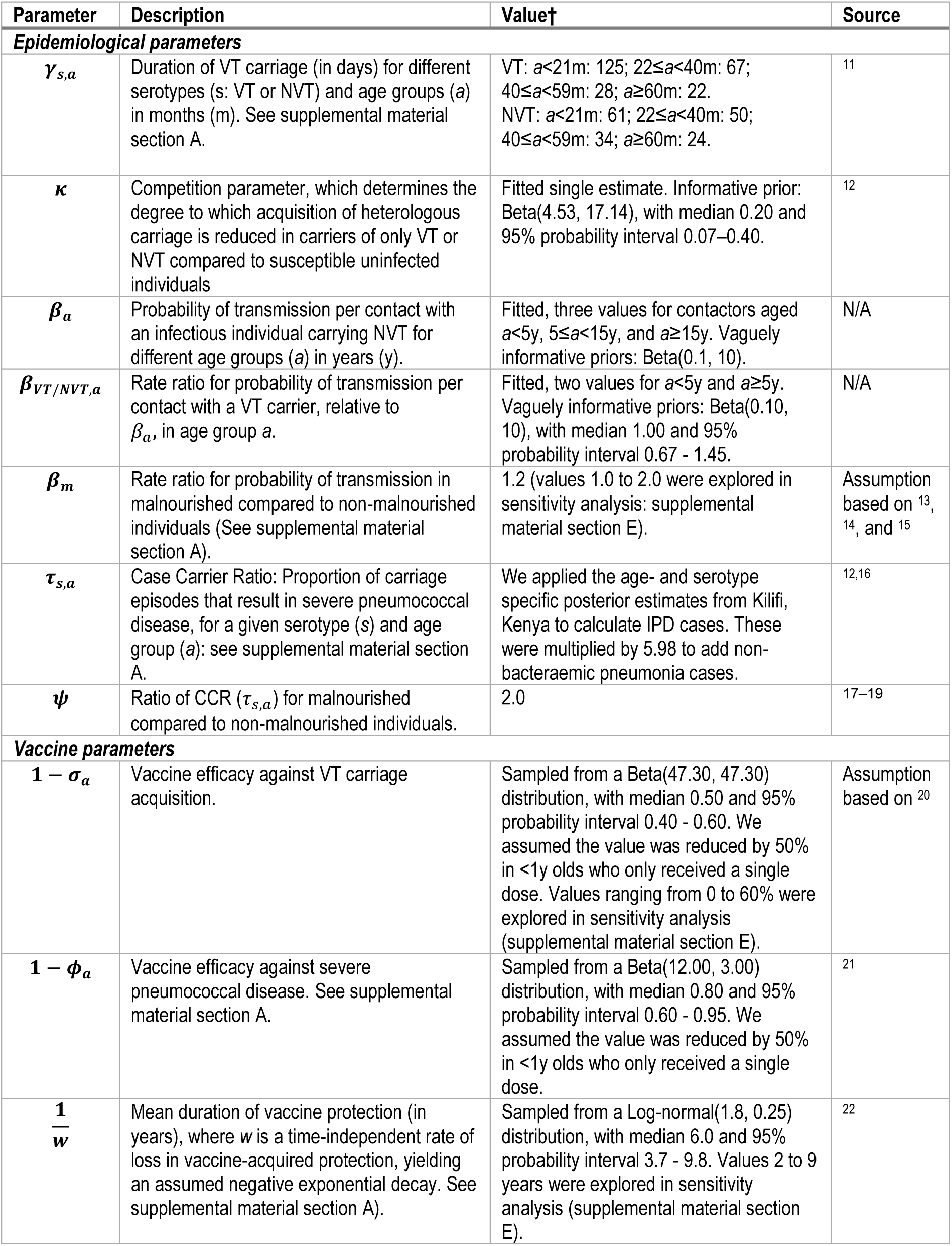
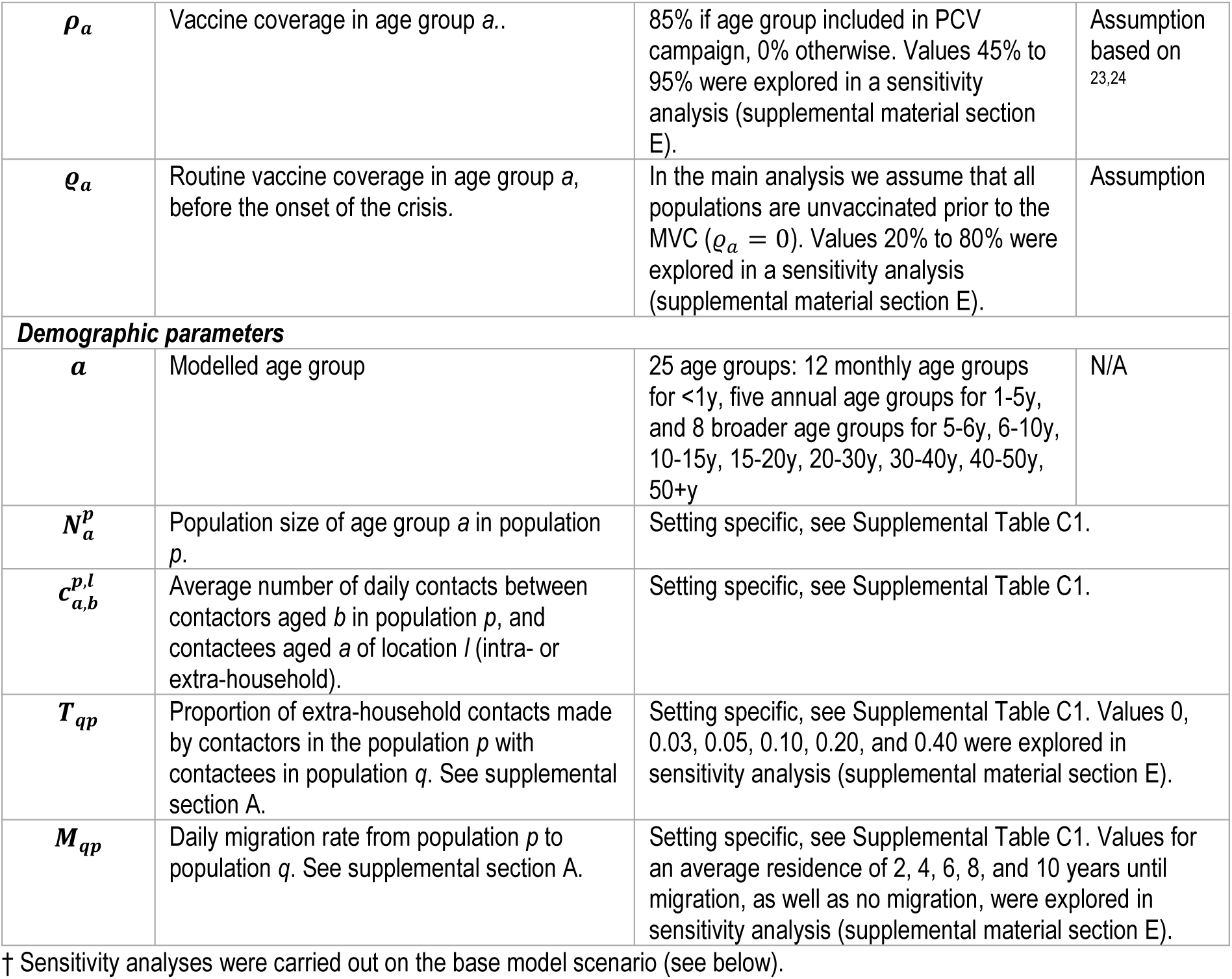
Overview of model parameters.

We modelled the proportion of people within each population, age group, and compartment, assuming a constant population size and age distribution, and ignoring deaths directly related to pneumococcal disease as they are likely to be numerically negligible for transmission. Equal and opposite rates of in- and out-migration (additional to travel-related contact) are applied to maintain a constant population size.

### Epidemiological assumptions

We applied age-specific duration of carriage estimates for VTs and NVTs estimated from a longitudinal study in Kilifi, Kenya.^11^ Malnutrition is an important risk factor for pneumococcal disease that is often highly prevalent in crisis-affected populations, and may affect the age-distribution of cases.^4^ We stratified populations by acute malnutrition status (not malnourished, versus moderate or severe acute malnutrition) to account for increased risk of pneumococcal transmission and disease in malnourished individuals, assuming 20% higher acquisition rates and a 100% higher invasive pneumococcal disease (IPD) risk.^14,18^ We calculated the incidence of IPD by applying VT- and NVT-specific case-carrier ratio (CCR) estimates from Kilifi, Kenya^12^ to the modelled incidence of carriage acquisitions. We assumed that every IPD case corresponded to an additional 4.98 cases of non-bacteraemic pneumococcal pneumonia, as estimated in Kenya^16^ (Table 1 and supplemental material section A).

### Vaccine protection

We assumed that two doses in infants and a single dose in those aged 1y and older would result in a direct vaccine efficacy (VE) against VT carriage acquisition of 50% (range 40-60%), a direct VE against VT pneumococcal disease of 80% (range 60-95%), and an average duration of vaccine protection of 6 years (range 4-10).^20–22^ We sampled from distributions around these estimates to incorporate their uncertainty in our results. We assumed that these values were halved for infants who only received a single dose of PCV.^20^

### Base scenario: Digaale IDP camp, Somaliland

To parameterize our transmission model, we previously conducted a cross-sectional survey in Digaale in 2019.^4,9,10^ The permanent IDP camp was established in 2013 following droughts and famines, and is located near the city of Hargeisa, the capital of Somaliland. It has an estimated 3000 people living across 700 shelters. We collected data on social contacts (a proxy for pneumococcal transmission pathways^25^), pneumococcal carriage prevalence, and demographic estimates including mortality, migration, and malnutrition rates (Supplemental Table C1). The survey, contact data, and carriage data are described in more detail elsewhere^9,10^. Ethical approval was granted by the Somaliland Ministry of Health Development, Directorate of Planning, Policy, and Strategic Information (2/13075/2019) and by the London School of Hygiene & Tropical Medicine (16577).

### Additional exemplar crisis scenarios

To account for a wider diversity of distinct crisis-affected settings, we modelled an additional three examplar populations: i) an acute-phase IDP camp based on the Bentiu protection of civilians (PoC) site in Unity State, South Sudan in 2015, which is substantially bigger and more crowded than Digaale, ii) an acute-phase urban setting with mixed IDP and host communities based on the city of Maiduguri in Borno State, North-East Nigeria in 2016, and iii) a protracted-phase rural setting based on Bambari town, Ouaka Prefecture, Central African Republic. For each setting, we identified and extracted data on population demographics, malnutrition, and migration. In the absence of other pneumococcal prevalence estimates, we assumed the same prevalence as in Digaale. More information about the exemplars and their parameterization are provided in supplemental material section C.

### Model fitting

We fitted five parameters for the age- and serotype-specific probability that contact between a susceptible and carrying individual resulted in transmission, and one competition parameter to scale these probabilities for acquisition of heterologous carriage.

We used a Markov chain Monte Carlo algorithm to fit our model to age-specific carriage estimates from Digaale in 2019, comparing the model-predicted steady state prevalence with the observed age-dependent prevalence in each S, VT, NVT, and B compartment. The model was refitted for each crisis exemplar to ensure similar carriage prevalence with different model parameters.

### Vaccination strategies

We modelled a range of MVCs with different age eligibility: offering PCVs to children aged 6 weeks to 11 months, and extending this to children aged up to 1, 4, 9 and 14y. We assumed a single-dose regimen in all age groups as our base case and assumed that every campaign would reach 85% coverage^23,24^ in the eligible age groups, consistent with observed PCV campaign coverage estimates in humanitarian settings.^23,24^

We compared the effectiveness of PCV campaigns by their modelled impact on VT carriage prevalence and on severe pneumococcal disease cases. We quantified the efficiency of different campaigns by calculating the average number of vaccine doses needed to prevent one case of severe pneumococcal disease (NVN), given the cumulative cases predicted in the no-vaccination scenario. We also assessed the incremental NVN between campaigns, i.e. the number of additional vaccine doses needed to prevent one additional case of severe pneumococcal disease, where each campaign is compared to the campaign with the next narrower age eligibility.

We assumed that all populations were PCV-naïve pre-MVC but conducted a sensitivity analysis to explore the effect of various baseline levels of routine vaccination coverage on model outputs (supplemental material section D).

### Sensitivity analyses

We assessed the sensitivity of our results against different parameter values for the i) number of vaccine doses, ii) vaccination coverage, iii) duration of vaccine protection, iv) VE against carriage, v) migration rate, vi) mixing with the host population, and vii) effect of malnutrition on transmission. We also compared our main model to one in which superinfection with the same serotype group is possible (supplemental section E).

### Forecasting global demand

To explore resource and potential stockpiling requirements for PCV use in humanitarian settings, we estimated the demand for the annual humanitarian PCV doses needed in low- and lower middle-income countries over a five-year horizon, if PCV campaigns would systematically be used. We assumed that a single MVC would be conducted in all currently displaced and newly displaced populations, but did not account for maintenance strategies after the initial campaign (supplemental material section F).

### Code availability

All analyses were implemented in R and C++. The model code and data can be found at https://github.com/kevinvzandvoort/espicc-metavax-pcv-humanitarian-crises.

## Results

Our model closely reproduced the age-specific pre-vaccination prevalence and distribution of VT and NVT serotypes observed in Digaale (supplemental material section B).

### Impact on VT prevalence

In our base case model of Digaale IDP camp, all strategies reduced VT prevalence compared to the no-vaccination baseline. Widening the age eligibility of the PCV campaign increased both the peak and duration of impact on VT carriage prevalence (Figure 2). For MVCs including children <1, <2, <5, <10, or <15y, peak impact on infant VT prevalence was reached on day 98 (88 – 110), 171 (137 – 395), 336 (248 – 445), 374 (287 – 475), and 386 (297 – 487) following the campaign, respectively. Infant VT carriage prevalence reduced from 56% (47 – 64) in the absence of vaccination to 53% (44 – 61), 51% (42 – 60), 45% (34 – 54), 38% (27 – 49), and 36% (24 – 47) at peak impact. Without subsequent vaccination, VT carriage prevalence returned to its pre-MVC baseline after about one year for the <1y and <2y strategies, while the effect was prolonged for more than three years for strategies using wider age eligibility. Estimates over different time periods are provided in Supplemental Table C1.

**Figure 2.**
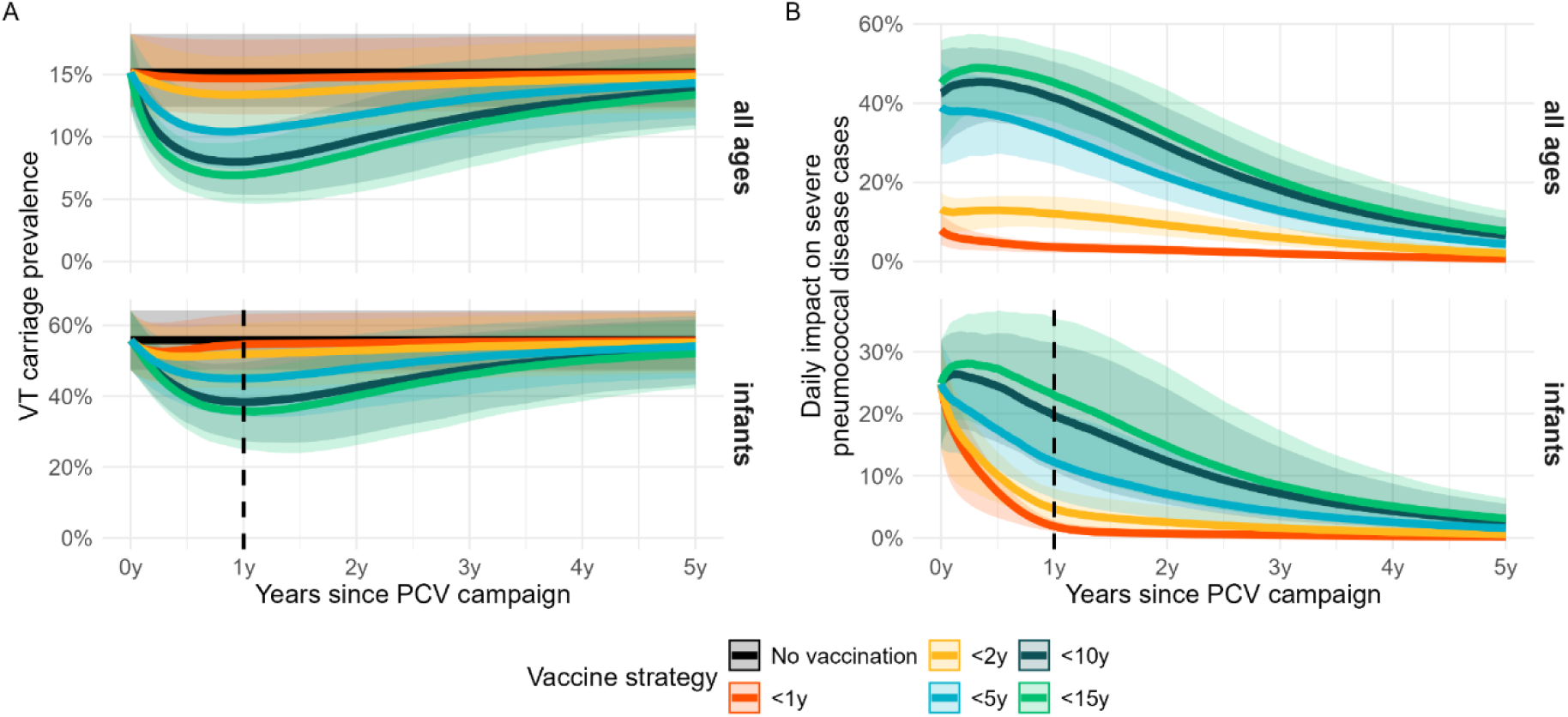
Impact of PCV mass vaccination campaigns on prevalence and incidence. A: VT carriage prevalence (VT + B) in all age groups and infants only. B: reduction in daily severe pneumococcal disease cases compared to no vaccination, in all age groups and infants only. In all plots, thick lines show the median estimates and shaded areas corresponding 95% credible intervals from 500 model runs. A dotted vertical line is plotted for infants 1 year after the PCV campaign: infants to the right of this line have been born after the PCV campaign and are thus all unvaccinated. Reductions in these birth cohorts result from indirect vaccine protection.

### Impact on disease

The cumulative number of severe pneumococcal disease cases prevented over a three-year period increased substantially with widening age eligibility (Table 2). Campaigns in infants and toddlers only had a small impact on disease, and only in the first year. The <5y campaign did prevent 13 (8 - 18) severe pneumococcal disease cases per 10,000 people, 27% (20 - 34) of all cases, while the <10 and <15y campaigns extended this to 16 (11 - 23) and 18 (13 - 24) cases prevented per 10,000 people, or 34% (26 – 43) and 38% (29 – 46) of all cases. Strategies vaccinating children up to 4y old or greater were able to generate sufficient indirect protection to substantially protect unvaccinated infants born after the MVC (Figure 2 and Table 2).

**Table 2.** Cumulative disease impact and efficiency of PCV campaigns over a three-year period.

| Age | Vaccine strategy | Cumulative disease impact per 10,000 people <sup>a</sup> | Number of vaccines needed to prevent one case (NVN) <sup>b</sup> |
| --- | --- | --- | --- |
| All ages | <1y | 2 (1 - 2, 4%) | 150 (105 - 249) |
|  | <2y | 5 (3 - 7, 10%) | 108 (77 - 166) |
|  | <5y | 13 (8 - 18, 27%) | 108 (77 - 162) |
|  | <10y | 16 (11 - 23, 34%) | 175 (125 - 250) |
|  | <15y | 18 (13 - 24, 38%) | 231 (167 - 326) |
| Infants | <1y | 16 (8 - 29, 4%) | 480 (258 - 979) |
|  | <2y | 25 (13 - 47, 6%) | 632 (339 - 1,300) |
|  | <5y | 50 (25 - 96, 11%) | 827 (434 - 1,700) |
|  | <10y | 76 (37 - 151, 16%) | 1,100 (570 - 2,300) |
|  | <15y | 88 (42 - 172, 19%) | 1,400 (724 - 3,000) |

- Estimates show the median absolute cumulative impact per 10,000 people. The 95% uncertainty intervals and median relative impact are provided in brackets. Estimates are taken from 500 model runs. - Estimates show the total number of PCV doses needed to prevent one case of severe pneumococcal disease. The 95% uncertainty intervals are provided in brackets. Estimates are taken from 500 model runs. Values greater than 1000 are rounded to the nearest hundred.

### Efficiency of PCV doses

The <5y campaign was the most efficient strategy over all time periods considered, with an average NVN of 108 (77 - 162) to prevent one case of severe pneumococcal disease over a three-year period (Table 2). As no PCVs were administered after the initial campaign, NVN values for all MCVs considered decreased with time as prevented cases accrue. While an <15y campaign was the least efficient strategy, its NVN value reduced from 549 (396 – 782) over a one-year period, to 231 (167 - 326) over a three-year period (Supplemental Table C2). NVN values were substantially higher when restricted to infant cases prevented, with decreasing efficiency as age eligibility widened.

The additional doses administered to children 1y old in an <2y campaign were 42% (31 – 53) more efficient in preventing severe pneumococcal disease cases than doses administered to infants in an <1y campaign (Figure 3). While the <5y campaign was the most efficient campaign overall, the additional doses administered to children 2-4y were 24% (13 - 38) less efficient in preventing pneumococcal disease than PCV doses administered to children 1y old in an <2y campaign. The additional doses administered to children aged 5-9 and 10-14y in <10y and <15y campaigns, were 281% (149 - 507) and 102% (71 - 139) less efficient compared to those administered to children aged 2-4 and 5-9y in campaigns with narrower age eligibility.

**Figure 3.**
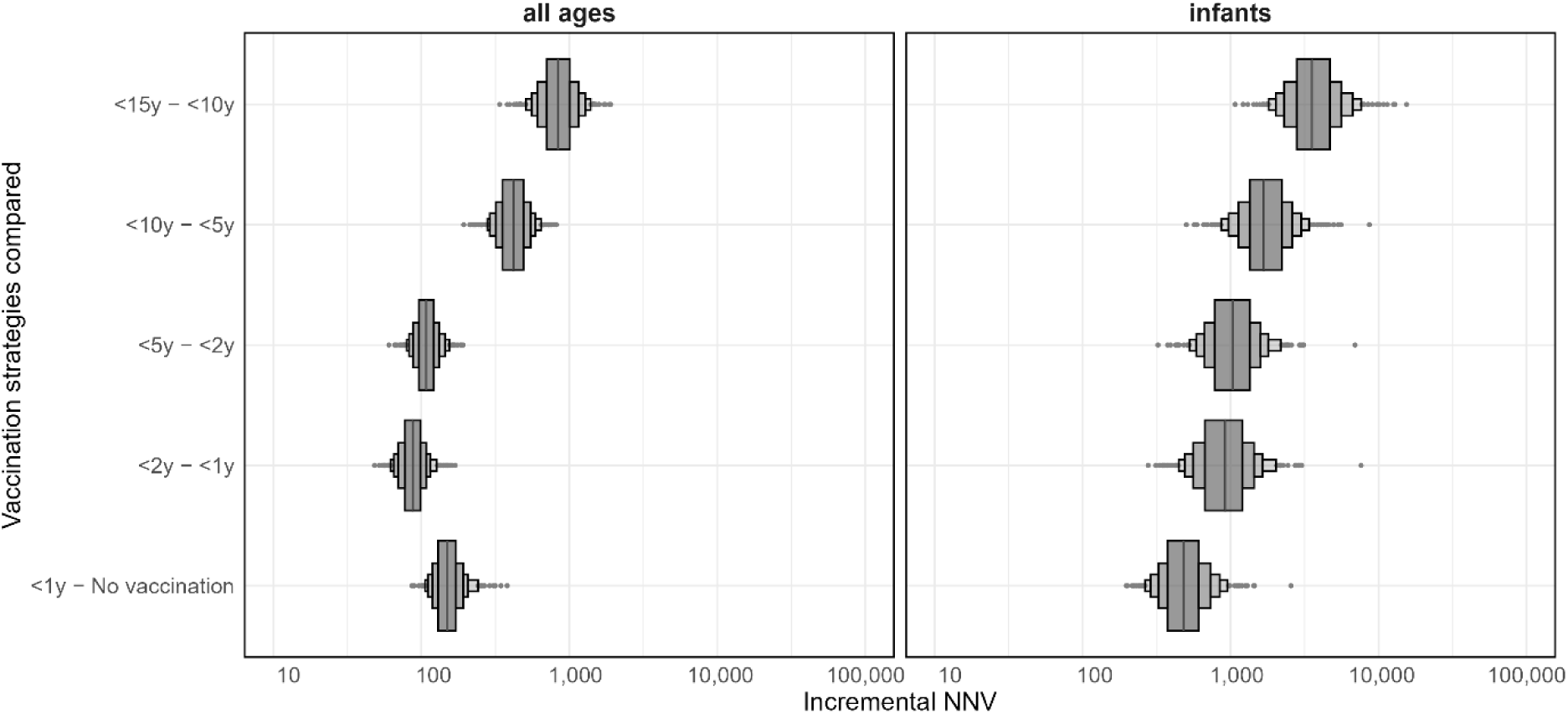
Incremental number of vaccines needed to prevent a single case of severe pneumococcal disease (NVN) over a three-year period. Estimates show the additional number of PCV doses needed to prevent one additional severe pneumococcal disease case over a three-year period, compared to a strategy with narrower age eligibility. Boxen plots show the distributions of the estimated incremental NVNs from 500 model iterations.

The <1y campaign had the lowest NVN to prevent one case of infant disease over a three-year period (480; 258 - 979). While wider age eligibility increased the impact on infant pneumococcal disease cases, the incremental efficiency per dose reduced by 88% (2 - 229) for doses administered to children 1y old, a further 14% (3 - 25) when vaccinating children 2-4y old, and a further 64% (29 - 122) and 110% (75 - 152) when vaccinating children 5-9 and 10-14y old.

### PCV campaign dosing schedule

Campaigns where infants received two doses were slightly more effective in reducing severe pneumococcal disease, particularly in infants (Figure 4). However, this increased impact was accompanied by reduced efficiency. With wider age eligibility, the relative differences in impact and NVN reduced compared to a single-dose campaign with the same age eligibility. A single-dose <5y campaign prevented a substantially larger proportion of cases (27%; 20 - 34) than a two-dose <2y campaign (12%; 8 - 16), while its NVN (108; 77 - 162) was lower than the two-dose <2y campaign (138; 100 - 200).

**Figure 4.**
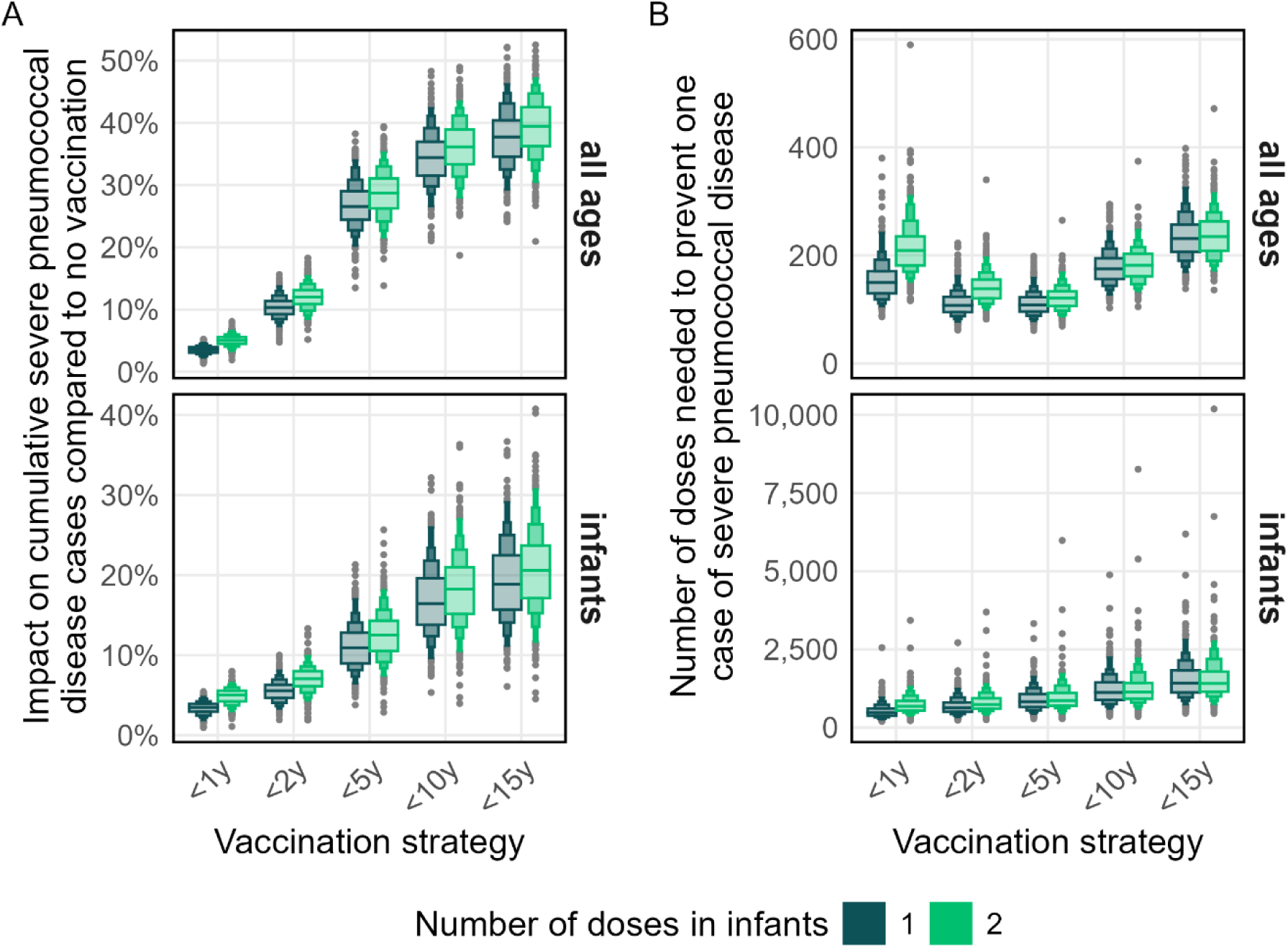
Comparing single-dose strategies to strategies where infants received two doses of PCV. A: impact on cumulative severe pneumococcal disease cases over a three-year period, in all age groups and infants only. B: NVN of vaccine strategies over a three-year period, in all age groups and infants only. Boxen plots show the distributions of the estimated values from 500 model iterations.

### Sensitivity analyses

The modelled impact on severe pneumococcal disease cases in Digaale was sensitive to the degree of interaction with the host population, migration rate, vaccine coverage, VE against transmission, and duration of vaccine protection, but not to the assumed increase in transmission in malnourished individuals (supplemental material section E). The sensitivity of model outputs to these parameter values was greater for the impact on infant cases than that in all age groups. In scenarios with decreased vaccination coverage, increased migration rates, or increased mixing between the displaced and host population, strategies with wider age eligibility were required to mitigate the loss in impact, at the expense of increased NVN compared to the base scenario. Increased vaccination coverage resulted in increased impact but not increased NVN for all MVCs.

While an alternative model allowing for same-serotype superinfections projected greater impact of all PCV campaigns, with longer duration and improved efficiency, it did not qualitatively alter the relative rank of each MVC strategy in terms of impact compared to the base model, nor did the NVN values of <1y, <2y, and <5y campaigns differ substantially (supplemental material section E).

### Other crisis exemplars

In all three exemplar settings, patterns were consistent with those estimated for Digaale (Figure 5), and the <5y campaign was consistently the most efficient campaign (Supplemental Table C2). The overall impact was most pronounced in Bambari and least in Maiduguri, highlighting the stark differences in their migration rates and interaction with an unvaccinated host population. In Maiduguri, an <10y campaign would be required to achieve similar impact to an <5y campaign in all other settings.

**Figure 5.**
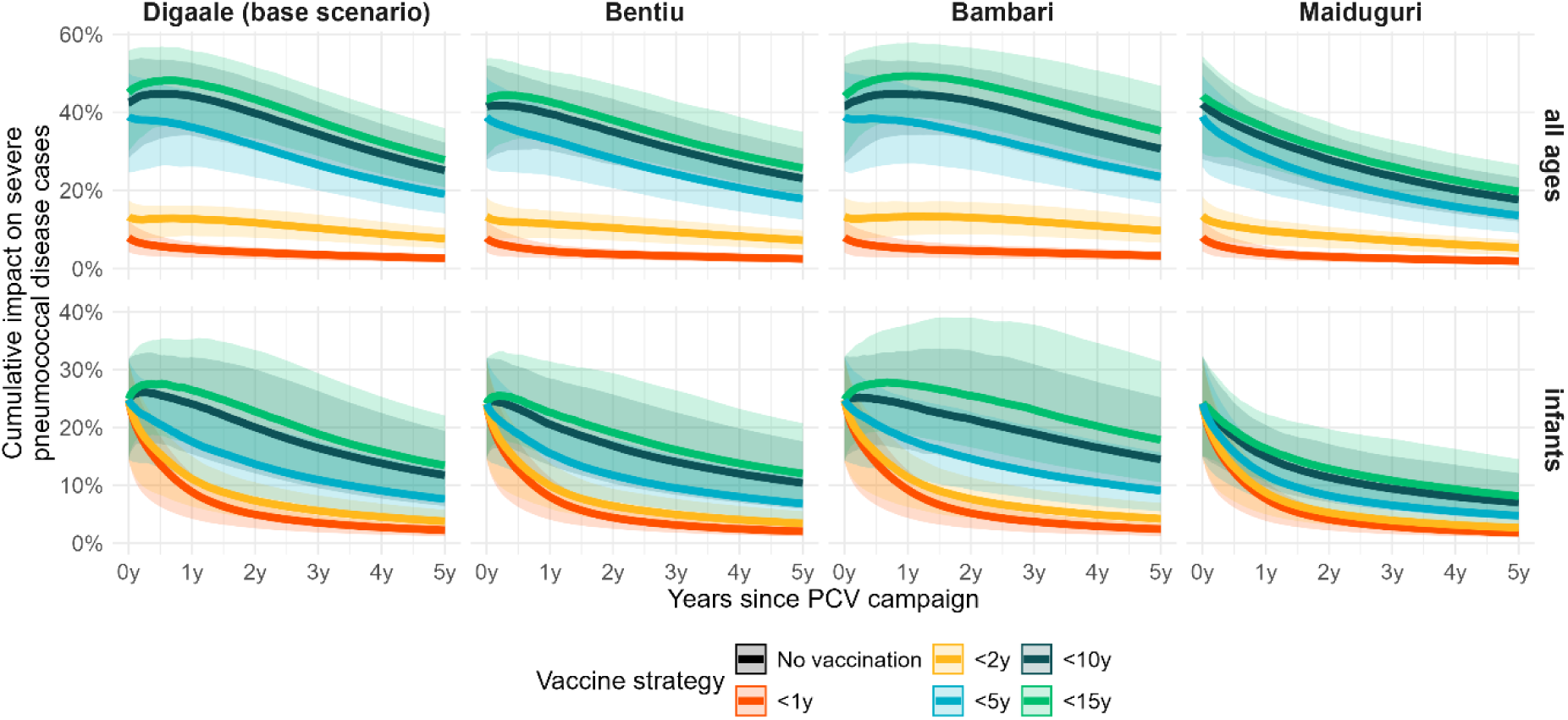
Cumulative impact of PCV campaigns in four settings. Plots show the cumulative impact over time in infants and all age groups for campaigns in Digaale, Bentiu, Bambari, and Maiduguri. In all plots, thick lines show the median estimates and shaded areas corresponding 95% credible intervals from 500 model iterations.

### Pre-campaign vaccination coverage

In our base scenario, we assumed a PCV-naïve population as observed in Somaliland. In a sensitivity analysis, increased PCV coverage pre-campaign resulted in lower baseline VT prevalence at the time of the campaign and thus reduced impact and efficiency of each campaign relative to these baseline levels, especially in the first year (Supplemental Material section D).

### Global humanitarian PCV demand

The number of people forcibly displaced people in low and lower middle-income countries has on average increased by 16 million people annually in the previous five years.^1^ If single-dose <5y or <10y PCV campaigns were implemented in all these populations, we estimate the humanitarian need for PCV to be 28 to 55 million doses over the next five years (supplemental material section F).

## Discussion

We found that single-dose PCV campaigns can be effective in reducing VT pneumococcal transmission and preventing pneumococcal disease in crisis-affected populations. The impact of a campaign with adequate age eligibility and vaccination coverage may be sustained for up to three years. While children <2y old are typically at highest risk of severe pneumococcal disease, the effectiveness of a campaign that only vaccinated this age group was low and short-lived. Campaigns that extend vaccination to children up to 4y or older achieved considerably higher and longer-lived impact, in part resulting from substantial herd immunity extending protection to unvaccinated children born or migrated into the population after the campaign. We previously found that children aged 2-5 and 6-14y were the main drivers of pneumococcal transmission in Digaale.^9^ Vaccinating these age groups is essential to maximize the impact and duration of a PCV campaign. Increased vaccination coverage resulted in greater impact without loss of efficiency of additional doses administered.

Campaigns that vaccinated children <5y old were consistently the most efficient use of PCV in all scenarios considered, although their overall impact was lower in settings that had i) higher rates of mixing with an unvaccinated host population, or ii) high migration rates. In such settings, wider age eligibility could partially mitigate this reduced impact, as suggested in the WHO Framework for Decision-Making on Vaccination in Humanitarian Emergencies^26^, albeit with reduced efficiency.

While administering two doses in infancy prevented more disease cases than a single dose, this strategy could be logistically challenging in many humanitarian crises. Our results suggest that allocating resources towards a more feasible strategy that vaccinates a larger group of children with a single dose may be more impactful and efficient.

We did not assess the cost-effectiveness of PCV strategies directly, but Gargano et al. assessed the cost-effectiveness of PCV10 immunization campaigns during humanitarian crises vaccinating children <1y in Somalia and <2y in a camp in South Sudan using static models.^23,27^ They found these PCV campaigns to be highly cost-effective using WHO cost-effectiveness thresholds. While results are not directly comparable, our results suggest that single-dose campaigns with extended age eligibility up to 4y are more efficient than those vaccinating <1y and <2y olds, and are thus likely cost-effective. Integration with other interventions, e.g. with measles in multi-antigen campaigns, may further improve cost-effectiveness. In humanitarian responses, measles campaigns typically vaccinate children up to 14y: applying the same age limit for PCV could be a pragmatic option that will, in addition, be robust to unforeseen demographic changes such as sudden large-scale in-migration, deteriorating nutritional status or further displacement placing the population in contact with less vaccinated groups.

There are several limitations to our analysis. First, we grouped pneumococcal serotypes as VT and NVT. This ignores differences between individual serotypes but is a common method used to effectively model PCV impact in other settings.^12,16^ We assumed a similar VE as has been estimated for other PCV products, but there is little evidence on the VE against VT transmission of reduced-dose PCV schedules, and none for Pneumosil. While we collected key data from the Digaale IDP camp, we had to assume similar pneumococcal carriage prevalence in other settings and extrapolate collected contact estimates. In the absence of pneumococcal disease data in Digaale or other humanitarian settings, we applied CCRs estimated in Kenya, a setting with similar prevalence and age distribution of pneumococcal carriage.^12^ While we adjusted these CCRs for differences in the prevalence of acute malnutrition, we do not know whether the presence of other risk factors may alter the virulence of VT and NVT serotypes in more fragile settings. Finally, in the absence of subsequent immunization activities or other changes, VT transmission will return to pre-vaccination levels. Alternative maintenance strategies need to be considered if routine immunization cannot be established within three years following an MVC, but we did not assess these here.

Respiratory diseases are a leading cause of mortality in crisis-affected populations, but pneumococcal vaccines remain underutilized.^3,28^ There is little empirical data on the effectiveness of PCV campaigns in humanitarian settings: a single-dose PCV campaign vaccinating children aged 1-9y was shown to be effective in reducing VT carriage prevalence in Niger, ^29^ and we are currently evaluating the effectiveness of an <5y campaign in the Digaale IDP camp. In the absence of comprehensive observed data, our results can support decision makers in deciding on the age range and dosing strategy for PCV campaigns. If such campaigns are to be implemented regularly, 40 to 64 million doses may be needed to protect crisis-affected populations over the next five years.

## Data Availability

All data produced in the present study are available upon reasonable request to the authors

## Acknowledgements

We like to thank all study participants who dedicated their time to the cross-sectional survey and the staff conducting the survey in Digaale. We also like to thank Médecins Sans Frontières for their participation in the inception of this work, and the MCRI Translational Microbiology Group for advice and laboratory testing of samples.

