## Supplemental Material for "Effectiveness of pneumococcal vaccination campaigns in humanitarian settings: a modelling study"

Analysis scripts and model code are available on GitHub:

https://github.com/kevinvzandvoort/espicc-metavax-pcv-humanitarian-crises. These can be used to conduct all simulations and analyses in both the main manuscript and the supplemental material.

### Section A. Transmission model

#### *Model structure and equations*

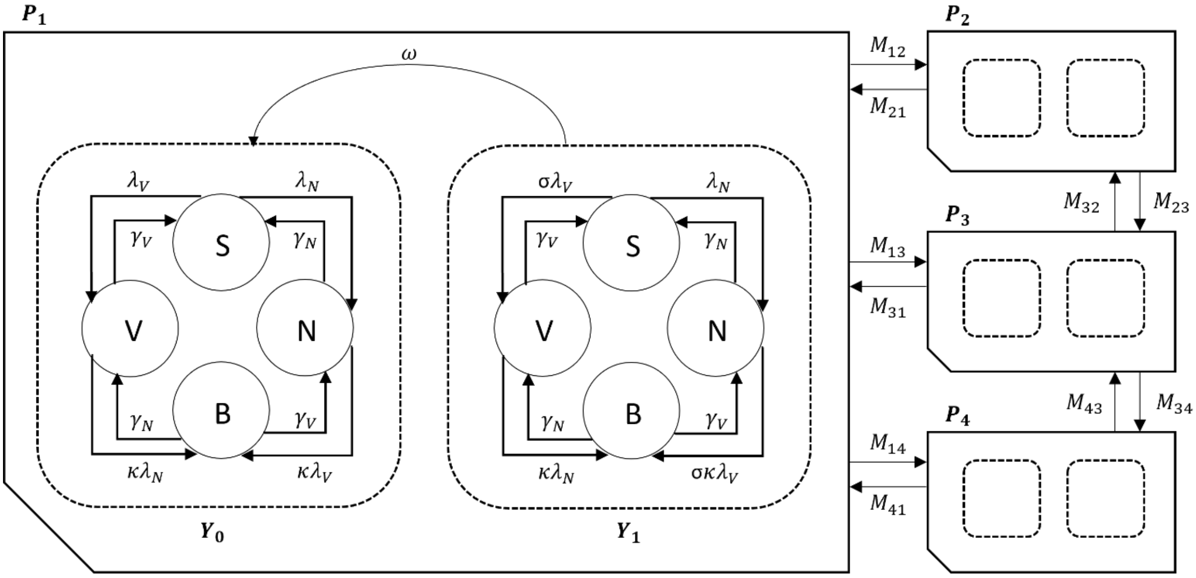

**Supplemental Figure A1. Compartmental model structure of the pneumococcal transmission model.** Pneumococcal serotypes defined as those included in the PCV vaccine (vaccine-type, VT), or not included in the vaccine (non-vaccine type, NVT). At any time $t$, Susceptible individuals (S) acquire serotypes in either group at rates $\lambda_V(t)$ and $\lambda_N(t)$, and immediately become infectious carriers themselves. Carriers of VTs or NVTs remain partially susceptible to infection with serotypes in the other group, and may develop a superinfection (B) at rates $\kappa\lambda_V(t)$ and $\kappa\lambda_N(t)$, where $\kappa$ represents competition between serotypes. Carriers are assumed to clear their infection of all serotypes in either group at rates $\gamma_V$ for VTs and $\gamma_N$ for NVTs. Individuals may be without vaccine derived protection ($Y_0$), or with full vaccine derived protection ($Y_1$). Those with vaccine protection acquire VT at a reduced rate $\sigma$, and lose their level of protection at rate $\omega$. Different populations $P_1$, $P_2$, and $P_3$, $P_4$ represent malnourished and non-malnourished strata of a displaced and host population. Individuals of population $p$ migrate to population $q$ at rate $M_{qp}$. Age groups, ageing, and transmission between vaccine protection strata, populations, and age groups are not shown. Note, Figure 1 in the main manuscript does not show the stratification of each population by malnutrition for improved clarity, which is why the number of populations in this figure differs.

#### *Modelled populations*

We constructed two model populations for every population in a scenario, to stratify these populations by the proportion with acute malnutrition. In our mathematical notation, we will refer to these as the model population (p) and real population (r). For example, in our base scenario in Digaale IDP camp, we created four model populations ($N_{\text{pop}} = 4$) to represent two real populations ($N_{\text{real}} = 2$): a non-malnourished (p=1) and malnourished (p=2) population for the displaced population living in Digaale IDP camp (r=1), and a non-malnourished (p=3) and malnourished (p=4) population for the host population in the city of Hargeisa (r=2). All populations (p and r) have the same number of age groups ($N_{\text{age}}$).

**Supplemental Table A1. Modelled populations in the base model.**

| Real population index (r) | Model population index (p) | Description |
|---|---|---|
| 1 | 1 | Digaale – non malnourished |
| 1 | 2 | Digaale – malnourished |
| 2 | 3 | Hargeisa – non malnourished |
| 2 | 4 | Hargeisa - malnourished |

We calculated the population size for age group *a* in model population p ($P_a^p$) by combining the population size for age group *a* in real population *r* ($R_a^r$) with the proportion of malnourished individuals in real population *r* ($\chi_r$)*:*

$$P_a^{2(r-1)} = R_a^r(1-\chi_r) \quad (1)$$

$$P_a^{2r} = R_a^r\chi_r \quad (2)$$

Every model population is further stratified in two vaccinated strata (Y; $N_{\text{vac}} = 2$), where y=0 indicates the unvaccinated stratum, and $y = 1$ the vaccinated stratum, and in four epidemiological compartments: S (Susceptible), V (VT carrier), N (NVT carrier) and B (carrier of both VT and NVT). The values for epidemiological compartment $X_a^{p,y}$, where $X \in \{S, V, N, B\}$ (i.e. $S_a^{p,y}$, $V_a^{p,y}$, $N_a^{p,y}$, $B_a^{p,y}$), then hold the proportion of people in age group *a* in population *p*, that are in compartment $X$ in vaccinated stratum *y*. We specifically model proportions, where all values for model compartments within a single age group in a model population sum to 1, i.e.

$$\sum_{y=0}^{N_{\text{vac}}}\left(S_a^{p,y} + V_a^{p,y} + N_a^{p,y} + B_a^{p,y}\right) = 1 \quad (3)$$

**Supplemental Table A2. Parameters used in the model equations.**

| Parameter | Description |
|---|---|
| $N_{\text{real}}$ | Number of ‘real populations’ (e.g., a displaced or host population) |
| $N_{\text{pop}}$ | Number of model populations |
| $N_{\text{age}}$ | Number of age groups in model |
| r | Index of real population |
| p, q | Index of model population |

| | |
|---|---|
| a,b | Index of age group |
| $R_a^r$ | Population size in age group a in real population r |
| $P_a^p$ | Population size in age group a in model population p |
| $\chi_r$ | Proportion acutely malnourished in real population r. |
| $N_{\text{vac}}$ | Number of vaccinated strata in model |
| y, z | Index of vaccinated stratum |
| t | Time |
| $X_a^{p,y}(t)$ | Value of epidemiological compartment at time t; proportion of age group a in population p that is in epidemiological compartment X in vaccinated stratum y, where $X \in \{S, V, N, B\}$ |
| $S_a^{p,y}(t)$ | Epidemiological compartment for susceptibles |
| $V_a^{p,y}(t)$ | Epidemiological compartment for VT carriers |
| $N_a^{p,y}(t)$ | Epidemiological compartment for NVT carriers |
| $B_a^{p,y}(t)$ | Epidemiological compartment for carriers of both VT and NVT |
| $\Delta D_{X_a^{p,y}}(t)$ | Change in the demography of compartment $X_a^{p,y}$ at time t |
| $\alpha_a$ | Daily rate at which individuals age to the next age stratum. |
| $l_a$ | Life expectancy for age group a, i.e. the number of years an individual is expected to remain in age group a. |
| $M_{qp}$ | Migration matrix: daily per capita rate at which individuals migrate out of model population p and into model population q |
| $J_a^{qp}$ | Daily per capita rate at which an individual in age group a in population p migrates into population q |
| $\mu_a^p$ | Daily rate at which individuals in age group a migrate out of model population p |
| $\omega_a^{p,y}$ | Daily rate at which vaccine protection wanes for individuals of age a in vaccinated stratum y in population p |
| $\Delta E_{X_a^{p,y}}(t)$ | Change in the epidemiology of compartment $X_a^{p,y}$ at time t |
| $\sigma_a^{p,y}$ | Relative risk (1 – vaccine efficacy) of acquiring VT serotypes compared to unvaccinated individuals for individuals of age a in vaccinated stratum y in population p |
| $\lambda_{V_a^p}(t)$ | Force of infection of VT on people in age group a in population p, at time t |
| $\lambda_{N_a^p}(t)$ | Force of infection of NVT on people in age group a in population p, at time t |
| $\gamma_{V_a}$ | Daily rate at which a VT carrier in age group a clears VT pneumococci |
| $\gamma_{N_a}$ | Daily rate at which an NVT carrier in age group a clears NVT pneumococci |
| $\kappa$ | Competition parameter |
| $\beta_a^{p,V}$ | Combined fitted values that adjust the force of infection of VT on people in age group a in population p |
| $\beta_a^{p,N}$ | Combined fitted values that adjust the force of infection of NVT on people in age group a in population p |
| $\beta_a$ | Probability of transmission per contact with an infectious individual carrying NVT for different age groups (a) |
| $\beta_{VT/NVT,a}$ | Rate ratio for probability of transmission with contacts carrying VT, compared to NVT in age group a. |
| $\beta_m$ | Rate ratio for probability of transmission in malnourished compared to non-malnourished individuals. |
| $c_{ba}^p$ | Average number of daily contacts made by a contactor in age group a in population p with contactees in age group b |
| $o^p$ | Proportion of daily contacts made by a contactor in population p that are made outside of the household |
| $T_{qp}$ | Travel matrix: proportion of extra-household contacts made by a contactor in population p with contactees in population q |

We separated our model equations in those that model the changes in demography between different strata, which is the same for each compartment $X$, and those that model the epidemiological transitions between the model compartments, which is the same for each stratum $X_a^{p,y}$.

*Demographic transitions*

We first model the change in the demography for each compartment $X_a^{p,y}$ as $\Delta D_{X_a^{p,y}}(t)$:

$$\begin{aligned}\Delta D_{X_a^{p,y}}(t) = &-\left(\alpha_a + \mu_a^p + \omega_a^{p,y} - \alpha_a\left(\mu_a^p + \omega_a^{p,y}\right) - \mu_a^p\omega_a^{p,y} - \alpha_a\mu_a^p\omega_a^{p,y}\right)X_a^{p,y}(t) \\ &+ \alpha_a\left(1 - \mu_a^p\right)\left(1 - \omega_{a-1}^{p,y}\right)X_{a-1}^{p,y}(t) \\ &+ \delta_{y,0}\left(1 - \mu_a^p\right)\sum_{z=1}^{N_{\text{vac}}}(1 - \alpha_a)\omega_a^{p,z}X_a^{p,z}(t) + \alpha_a\omega_a^{p,z}X_{a-1}^{p,z}(t) \\ &+ \sum_{q=1}^{N_{\text{pop}}} J_a^{qp}\left((1 - \alpha_a)\left(1 - \omega_a^{q,y}\right)X_a^{q,y}(t) + \alpha_a\left(1 - \omega_{a-1}^{q,y}\right)X_{a-1}^{q,y}(t)\right) \\ &+ \delta_{y,0}\sum_{q=1}^{N_{\text{pop}}}\sum_{z=1}^{N_{\text{vac}}} J_a^{qp}\left((1 - \alpha_a)\omega_a^{q,z}X_a^{q,z}(t) + \alpha_a\omega_a^{q,z}X_{a-1}^{q,z}(t)\right)\end{aligned} \quad 4$$

- $\alpha_a$ is the daily rate at which people age out of age group a. We apply a negative exponential rate to incorporate ageing, calculated as

$$\alpha_a = \frac{1}{365.25 l_a} \quad 5$$

- where $l_a$ is the life expectancy or size for age group *a* in years, i.e. the number of years an individual should remain in age group *a*. For instance, if an individual in age group *a* is 2-3mo: $l_a = \frac{1}{12}$, while if they are 5-9yo: $l_a = 4$.
- $\mu_a^p$ is the daily rate at which people of age a migrate out of population *p*. It is calculated as

$$\mu_a^p = \sum_{q=1}^{N_{\text{pop}}} J_a^{qp} \quad 6$$

- where $J_a^{qp}$is the daily per capita migration rate at which individual in age group a migrate from population *p* to population *q*.
- $\omega_a^{q,z}$ is the rate at which vaccinated individuals lose vaccine protection and return to compartments in vaccinated stratum $y = 0$.
- $\delta_{y,0}$ denotes the Kronecker delta function which evaluates to 1 when $y = 0$, and evaluates to 0 for all other values of $y$.

The different elements of equation 4 can then be read as

- All those who leave compartment $X_a^{p,y}$ due to ageing, migration, or loss of vaccine protection, accounting for competing rates to keep population sizes consistent.
- Those who age into compartment $X_a^{p,y}$ from $X_{a-1}^{p,y}$. Note that newborns are a special case: when $X_a^{p,y} = S_0^{p,y}$, this term changes to $\alpha_a\left(1-\mu_a^p\right)\left(1-\omega_{a-1}^{p,y}\right)$, while it changes to 0 when $X_a^{p,y} \neq S_0^{p,y}$(i.e. all newborns are born susceptible).
- Those from model population p who move back into compartment $X_a^{p,y}$due to loss of vaccine protection (only when $y = 0$).
- Those from other model populations q who migrate into compartment $X_a^{p,y}$.
- And those from other model populations q who lose their vaccine protection and simultaneously migrate into compartment $X_a^{p,y}$ (only when $y = 0$).

Note that the $\alpha_a$ rates applied to the $X_{a-1}^{p,y}$ compartments and the $J_a^{qp}$ rates applied to the $X_a^{q,y}$ compartments are not typos; they adjust for any differences between $l_a$ and $l_{a-1}$ or between $P_a^p$and $P_a^q$, and thereby implicitly reweigh the values for compartment $X_a^{p,y}$.

*Epidemiological transitions*

With the demographic changes incorporated, we can then model the change in the epidemiology for each compartment $X_a^{p,y}$ as $\Delta E_{X_a^{p,y}}(t)$. To do so, we want to use the updated values accounting for the changes in demography:

$$\overline{X}_a^{p,y}(t) = X_a^{p,y}(t) + \Delta D_{X_a^{p,y}}(t) \quad 7$$

We can then calculate the epidemiological transitions. For ease of reading, we have omitted the time notations in the following set of equations. We have also given the notations for the age group, population, and vaccinated stratum a lighter colour:

$$\Delta E_{S_a^{p,y}}(t) = -\left(\sigma_a^{p,y}\lambda_{V_a}^{p} + \lambda_{N_a}^{p}\right)\overline{S}_a^{p,y} + \gamma_{V_a}\overline{V}_a^{p,y} + \gamma_{N_a}\overline{N}_a^{p,y}$$

$$\Delta E_{V_a^{p,y}}(t) = \sigma_a^{p,y}\lambda_{V_a}^{p}\overline{S}_a^{p,y} - \left(\kappa\lambda_{N_a}^{p} + \gamma_{V_a}\right)\overline{V}_a^{p,y} + \gamma_{N_a}\overline{B}_a^{p,y}$$

$$\Delta E_{N_a^{p,y}}(t) = \lambda_{N_a}^{p}\overline{S}_a^{p,y} - \left(\sigma_a^{p,y}\kappa\lambda_{V_a}^{p} + \gamma_{N_a}\right)\overline{N}_a^{p,y} + \gamma_{V_a}\overline{B}_a^{p,y}$$

$$\Delta E_{B_a^{p,y}}(t) = \kappa\lambda_{N_a}^{p}\overline{V}_a^{p,y} + \sigma_a^{p,y}\kappa\lambda_{V_a}^{p}\overline{N}_a^{p,y} - \left(\gamma_{V_a} + \gamma_{N_a}\right)\overline{B}_a^{p,y}$$

$$\lambda_{V_a}^{p} = \beta_a^{p,V}\sum_{b=1}^{N_{\text{age}}}\sum_{q=1}^{N_{\text{pop}}}\left(c_{ba}^{p}\left(\delta_{q,p}(1-o^p) + o^pT_{qp}\right)\sum_{y=0}^{N_{\text{vac}}}\left(\overline{V}_b^{q,y} + \overline{B}_b^{q,y}\right)\right)$$

$$\lambda_{N_a}^{p} = \beta_a^{p,N}\sum_{b=1}^{N_{\text{age}}}\sum_{q=1}^{N_{\text{pop}}}\left(c_{ba}^{p}\left(\delta_{q,p}(1-o^p) + o^pT_{qp}\right)\sum_{y=0}^{N_{\text{vac}}}\left(\overline{N}_b^{q,y} + \overline{B}_b^{q,y}\right)\right) \quad 8$$

- $\sigma_a^{p,y}$ is the relative rate (1 – vaccine efficacy against carriage) that scales the force of infection for individuals in age group a, vaccine stratum y, and population p.
- $\gamma_{V_a}$ and $\gamma_{N_a}$ are the rates at which VT and NVT carriers of age a clear their infection of that serotype.
- $\kappa$ is the competition parameter and determines the degree by which acquisition of heterologous carriage is reduced in carriers of only VT or NVT compared to susceptible individuals.

- $\beta_a^{p,V}$ and $\beta_a^{p,N}$ are calculated from fitted parameters. They are the probabilities of transmission per contact by an individual in age group a with infectious individuals carrying VT or NVT respectively. They are calculated as:

$$\beta_a^{p,N} = \begin{cases} \beta_a \beta_m, & p \bmod 2 = 0 \\ \beta_a, & p \bmod 2 \neq 0 \end{cases} \qquad 9$$

  - where $\beta_a$ is the fitted probability of transmission per contact by an individual in age group a with infectious individual carrying NVT, and $\beta_m$is the rate ratio for this probability of transmission for malnourished compared to non-malnourished individuals;

  and

$$\beta_a^{p,V} = \beta_a^{p,N} \beta_{VT/NVT,a} \qquad 10$$

  - where $\beta_{VT/NVT,a}$ is the fitted rate ratio for the probability of transmission with contacts carrying VT, compared to NVT, in age group *a*.

  See section B for more details about fitted model parameters.
- $c_{ba}^p$ are the average number of daily contacts made by an individual in age group *a* in population *p*, with contactees in age group *b*. Within age groups, contact rates are assumed to be homogenous irrespective of the individual's vaccine protection or malnourished stratum.
- $o^p$ is the proportion of contacts made by contactors in population *p* that take place outside of the household.
- $\delta_{q,p}$ denotes the Kronecker delta function which evaluates to 1 when $q = p$, and evaluates to 0 for all other values of *q*.
- $T_{qp}$ is the proportion of extra-household contacts that are made by a contactor in population *p* with contactees in population *q*.

We then combine the demographic and epidemiological transitions, and calculate the change for each model compartment at time *t* as:

$$\frac{dX_a^{p,y}}{dt} = \Delta D_{X_a^{p,y}}(t) + \Delta E_{X_a^{p,y}}(t) \quad 11$$

#### *Model parameter values*

##### *Extrapolating carriage to disease*

For each age group and population, we first calculated the incidence of all VT and NVT acquisitions at time *t* as:

$$\text{acquisitions}_{\text{VT}_a}^{p,y}(t) = P_a^p\left(\sigma_a^{p,y}\lambda_{V_a}^{p}(t)\overline{S}_a^{p,y}(t) + \sigma_a^{p,y}\kappa\lambda_{V_a}^{p}(t)\overline{N}_a^{p,y}(t)\right)$$

$$\text{acquisitions}_{\text{NVT}_a}^{p,y}(t) = P_a^p\left(\lambda_{N_a}^{p}(t)\overline{S}_a^{p,y}(t) + \kappa\lambda_{N_a}^{p}(t)\overline{V}_a^{p,y}(t)\right) \quad 12$$

We then applied age- and serotype specific case-carrier ratios (CCR) sampled from a posterior distribution estimated in Kilifi, Kenya by Flasche et al.[1].

We assumed that CCRs in malnourished individuals were increased by rate-ratio $\varphi$ compared to non-malnourished individuals. To also account for a relative increase in the acquisition of pneumococci in malnourished individuals, $\beta_m$, we calculated $\varphi$ so that the overall combined risk of IPD (increased risk in acquisition and disease) in malnourished individuals was increased by rate-ratio $\tau$ compared to non-malnourished individuals. as:

$$\varphi = \frac{\tau}{\beta_m} \quad 13$$

We then calculated the CCR for serotype *s* and age group *a* in non-malnourished individuals ($\text{CCR}_a^{s,0}$) as:

$$\text{CCR}_a^{s,0} = \frac{\text{CCR}_a^{s}}{\chi_0(\varphi - 1) + 1} \quad 14$$

Where $\text{CCR}_a^{s}$ is a sampled CCR value (combined for malnourished and non-malnourished individuals) for serotype s (VT or NVT) and age group *a*, and $\chi_0$ is the proportion of the population in Kilifi ($r = 0$) that was malnourished. In the baseline scenario, we assume that $\tau = 2.0$[2–4] and $\beta_m = 1.2$[5–7]. Furthermore, we assume that $\chi_0 = 0.182$[8].

The CCR for malnourished individuals was then calculated as:

$$\text{CCR}_a^{s,1} = \varphi \text{CCR}_a^{s,0} \qquad 15$$

These adjusted CCRs was then applied to each respective population, effectively scaling the estimated CCRs from Kilifi to settings with different rates of malnutrition.

*Vaccine efficacy against disease*

PCVs protect against both transmission and disease. Like we did for the rate-ratio $\varphi$ for malnourished individuals, we calculate the marginal vaccine efficacy (VE) against disease $(1 - \theta_a^{p,y})$ used in our model, given the VE against carriage $(1 - \sigma_a^{p,y})$ and the combined VE against carriage and disease $(1 - \phi_a^{p,y})$, which is often reported in studies of VE:

$$1 - \theta_a^{p,y} = 1 - \frac{\phi_a^{p,y}}{\sigma_a^{p,y}} \qquad 16$$

*IPD cases*

Total IPD cases at time *t* for age group *a* in vaccine stratum *y* in population *p* were then calculated as:

$$\text{cases}_{\text{VT}}{}_a^p(t) = \begin{cases} \text{acquisitions}_{\text{VT}}{}_a^{p,y}(t)\text{CCR}_a^{V,1}\theta_a^{p,y}, & p \bmod 2 = 0 \\ \text{acquisitions}_{\text{VT}}{}_a^{p,y}(t)\text{CCR}_a^{V,0}\theta_a^{p,y}, & p \bmod 2 \neq 0 \end{cases}$$

$$\text{cases}_{\text{NVT}}{}_a^p(t) = \begin{cases} \text{acquisitions}_{\text{NVT}}{}_a^{p,y}(t)\text{CCR}_a^{N,1}, & p \bmod 2 = 0 \\ \text{acquisitions}_{\text{NVT}}{}_a^{p,y}(t)\text{CCR}_a^{N,0}, & p \bmod 2 \neq 0 \end{cases} \qquad 17$$

In Kenya, it has been estimated that 4.98 cases of non-bacteraemic pneumonia are prevented for every case of IPD prevented.[9,10] We followed Ojal et al and calculated the total number of severe pneumococcal disease cases (the total number of IPD and non-bacteraemic pneumonia cases) as (1 + 4.98) times the estimated number of IPD cases.[11]

*Duration of carriage*

Lipsitch et al. estimated the age- and serotype-specific duration of carriage in children 0-22mo, 22-41mo, and 41-59mo from longitudinal data collected in Kenyan children.[12] Age-specific estimates for the 14 most common serotypes were provided in Figure 3 of their paper. We extracted these data and fitted negative binomial distributions to each set of values to recover their distributions.

For each serotype *s*, in each age *a*, we assume that the duration of carriage can be described by a negative binomial distribution with mean $\mu_{s,a}$ and dispersion parameter $k_{s,a}$.

$$d_{s,a} \sim \text{nbinom}(\mu_{s,a}, k_{s,a}) \quad (18)$$

We parameterized the mean value for $d_{s,a}$ as $\mu_{s,a} = y_{s,a,0.5}$, where $y_{s,a,0.5}$ is the extracted median estimate for serotype *s* in dataset *a.* We then optimized the value for $k_{s,a}$ using the *optimize* function in base *R* to minimize $g_{(d_{s,a})}$: the log-least squares of the difference between the extracted boundaries of the 95% confidence interval for $d_{s,a}$: $y_{s,a,0.025}$ and $y_{s,a,0.975}$, and corresponding quantile distribution for the negative binomial distribution evaluated at 0.025 and 0.975. We took the log of the values to minimize the large difference in scale between the lower and upper boundary value.

$$g(\sigma_{s,a}) = \log\left(\left(y_{s,a,0.025} - Q_{s,a}(0.025)\right)^2 + \left(y_{s,a,0.975} - Q_{s,a}(0.975)\right)^2\right) \quad (19)$$

Where $Q_{s,a}(p)$ is the quantile distribution of the negative binomial distribution used to describe $d_{s,a}$ evaluated at *p.*

Supplemental Figure A2 shows the fitted negative binomial distributions against the extracted values estimated by Lipsitch et al. The 95% quantile values of the fitted distributions were in broad agreement with the reported 95% confidence intervals, with only some minor discrepancies at primarily the lower tails of some distributions.

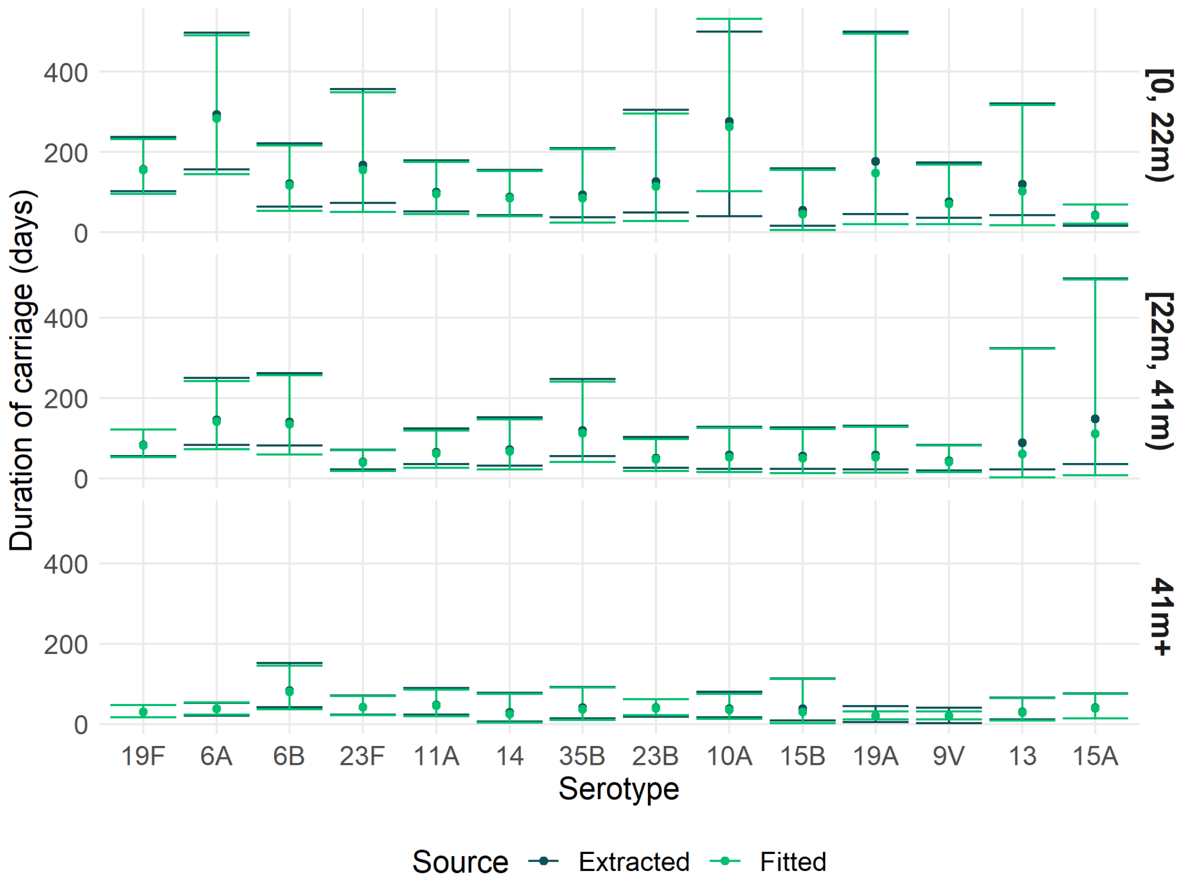

**Supplemental Figure A2.** Fitted distributions of age- and serotype-specific duration of carriage.

We then estimated 6 parameters independently to pool the average duration of carriage for VT and NVT in the three age groups, accounting for the uncertainty in the underlying distributions. We did so by finding the value of $\overline{\mu}_{\overline{s},a}$ that maximized the summed negative log-density of all fitted distributions $d_{s,a}$ maximized by the *optim* function, where the serotype was a VT or NVT (based on their inclusion in the Pneumosil vaccine).

The best fitting values for $\overline{\mu}_{\overline{s},a}$, for each age group and serotype group are described in Supplemental Table A3 and plotted against the serotype-specific distributions in Supplemental Figure A3.

For the duration of carriage in older people, we followed Flasche et al[1] and assumed that these were 36% lower for people aged 60m+ compared to those aged 24-59m, as estimated by Melegaro et al[13].

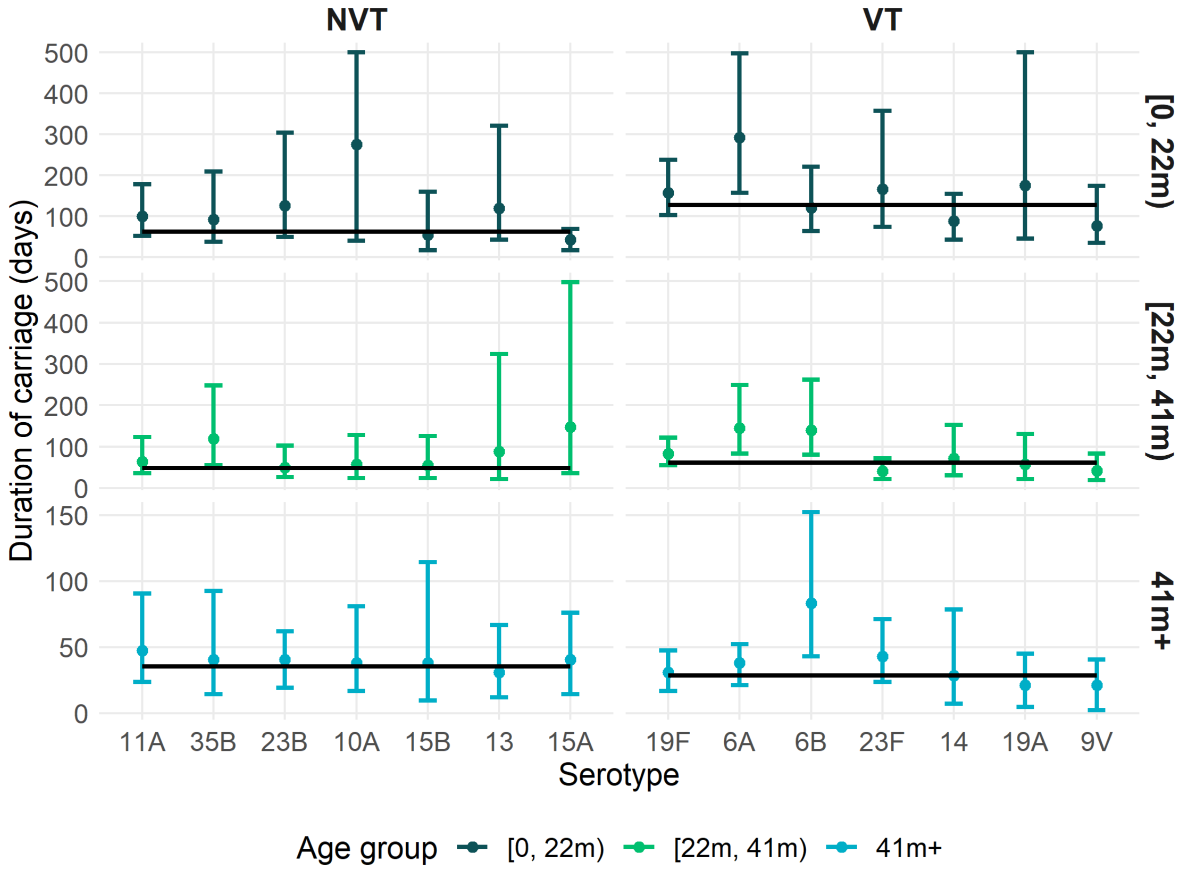

**Supplemental Figure A3.** Pool estimates of the duration of carriage for VT and NVT by age.

**Supplemental Table A3. Estimated duration of carriage (days) for VT and NVT serotypes by age.**

| Age group | VT | NVT |
|---|---|---|
| [0, 22m) | 125 | 61 |
| [22m, 41m) | 67 | 50 |
| [41m, 59m) | 28 | 34 |
| ≥ 60m | 22 | 24 |

*Transmission between populations*

We used a travel matrix to inform extra-household contacts and thus transmission between populations. For the displaced populations (r=1, p≤2), we assumed the same values for all age groups, informed by the value in the travel matrix ($T_{qp}$). These values were themselves calculated as the proportion of extra-household contacts made by an individual in model population *p* with individuals in real population *r*, multiplied by the proportion of real population *s* that is in model population *q*. The latter implicitly assumes that mixing between malnourished and non-malnourished individuals is at random.

$$\mathrm{T}_a^{qp} = \begin{cases} \dfrac{\sum_{b=1}^{N_{\text{age}}} P_b^q}{\sum_{b=1}^{N_{\text{age}}} R_b^{\lceil q/2 \rceil}} T_{qp}, & p \leq 2 \\ \dfrac{\sum_{b=1}^{N_{\text{age}}} P_b^p}{\sum_{b=1}^{N_{\text{age}}} R_b^2} T_{pq} \dfrac{o^q \sum_{b=1}^{N_{\text{age}}} P_b^q c_{ab}^q}{o^p P_a^p \sum_{b=1}^{N_{\text{age}}} c_{ba}^p}, & p > 2 \end{cases} \qquad 20$$

*Migration between populations*

For the displaced populations (r=1, p≤2) we assume the same migration rate for all age groups, informed by the value in the migration matrix ($M_{qp}$). For the host populations (r=2, p>2), the migration rate was calculated by the values in the displaced populations, adjusted for any difference in the age distributions and population sizes to ensure a fixed population size in all $R_a^r$:

$$J_a^{qp} = \begin{cases} M_{qp}, & p \leq 2 \\ M_{pq} \dfrac{P_a^q}{P_a^p}, & p > 2 \end{cases} \qquad 21$$

Individuals in the malnourished stratum in the displaced population only migrate to the malnourished stratum in the host population, and vice versa.

*Extrapolating contact estimates to other settings*

As we only had observed contact estimates for Digaale, we adjust the observed contact matrices to other settings as follows:

Matrix $\boldsymbol{C_{pr}}$ is a contact matrix in population *p* and relationship *r*, where value $c_{pr,ij}$ is the average number of daily contacts between a contactor in population *p* aged *i*, and contactees aged *j* with relationship *r*.

We assume that $c_{pr,ij}$ can be split in three elements:

$$c_{pr,ij} = h_{p,j} q_{pr,ij} d_{pr,i} \quad (22)$$

Here, value $h_{p,j}$ is the expected proportion of all contacts made by a contactor in population *p* aged *i*, that are with contactees aged *j*, if mixing would be completely homogeneous and at random. It is the same for all contactor age groups *i*, and equal to the proportion of people in population *p* who are of age *j*, i.e. $h_{p,j} = \frac{n_{p,j}}{\sum_{x=1}^{a} n_{p,x}}$ where $n_{p,j}$ is the total number of people in population *p* who are of age *j*, and *a* is the total number of age groups.

Value $q_{pr,ij}$ is the relative assortativity of contacts between contactors in population *p* aged *i*, and contactees aged *j* with relationship *r*. It is calculated as the relative difference between the real and expected (assuming homogeneous mixing) proportion of contacts between contactors in population *p* aged *i*, and contactees aged *j*, i.e. $q_{pr,ij} = \frac{\left(\frac{c_{pr,ij}}{\sum_{x=1}^{a} c_{pr,ix}}\right)}{h_{p,j}}$. Values between 0 and 1 would represent age-dissasortative mixing, a value of 1 would represent perfect homogeneous mixing, and values greater than 1 would represent age-assortative mixing.

Finally, value $d_{pr,i}$ represents the total average number of daily contacts made by a contactor in population *p* aged *i*, made with all contactees aged *j* and relationship *r*, i.e. $d_{pr,i} = \sum_{x=1}^{a} c_{pr,ix}$

To extrapolate the estimated contact rates from Digaale (p=0) to populations in other settings, without any additional information, we assumed that $q_{pr,ij}$and $d_{0r,i}$ remain the same as in Digaale, but update $h_{p,j}$ to the respective values for population *p*, and calculate the new contact rates as

$$c_{pr,ij} = h_{p,j} q_{0r,ij} d_{0r,i} \qquad 23$$

Note that we can rewrite this expression as:

$$\begin{aligned} c_{pr,ij} &= h_{p,j} q_{0r,ij} d_{0r,i} \\ &= h_{p,j} \frac{\left(\frac{c_{0r,ij}}{\sum_{x=1}^{a} c_{0r,ix}}\right)}{h_{0,j}} \sum_{x=1}^{a} c_{0r,ix} \\ &= c_{0r,ij} \frac{h_{p,j}}{h_{0,j}} \end{aligned} \qquad 24$$

It is worth noting that, as matrix $\boldsymbol{C_{0r}}$ was adjusted for reciprocity of contacts so that the total number of contacts made between contactors/contactees and contactees/contactors aged *i* and *j* with relationship *r* are equal (i.e. $\boldsymbol{c_{0r,ij} n_{0,i}} = \boldsymbol{c_{0r,ji} n_{0,j}}$), the same applies for the adjusted contact rates in population *p*:

$$\begin{gathered} c_{pr,ij} n_{p,i} = c_{pr,ji} n_{p,j} \\ c_{0r,ij} \frac{h_{p,j}}{h_{0,j}} n_{p,i} = c_{0r,ji} \frac{h_{p,i}}{h_{0,i}} n_{p,j} \\ c_{0r,ij} \frac{n_{p,j} \sum_{x=1}^{a} n_{0,x}}{n_{0,j} \sum_{x=1}^{a} n_{p,x}} n_{p,i} = c_{0r,ji} \frac{n_{p,i} \sum_{x=1}^{a} n_{0,x}}{n_{0,i} \sum_{x=1}^{a} n_{p,x}} n_{p,j} \\ \frac{c_{0r,ij}}{n_{0,j}} = \frac{c_{0r,ji}}{n_{0,i}} \\ c_{0r,ij} n_{0,i} = c_{0r,ji} n_{0,j} \end{gathered} \qquad 25$$

Therefore, no further adjustments for reciprocity of contacts need to be made.

We adjust the intra- ($\boldsymbol{C_{p0}}$) and extra-household ($\boldsymbol{C_{p1}}$) contact matrix separately. We assume that the total number of extra-household contacts is the same as observed in Digaale ($d_{p1,i} = d_{01,i}$):

$$c_{p1,ij} = c_{01,ij} \frac{h_{p,j}}{h_{0,j}} \tag{26}$$

We assume that the total number of intra-household contacts is proportional to the average household size in each setting ($d_{p0,i} = d_{00,i} \frac{s_p}{s_0}$), and calculate intra-household contacts as:

$$c_{p0,ij} = c_{00,ij} \frac{h_{p,j}}{h_{0,j}} \frac{s_p}{s_0} \tag{27}$$

$$d_{p0,i} = d_{00,i} \frac{s_p}{s_0} \tag{28}$$

Where $s_p$ is the average household size in population *p*. Note that this implicitly assumes that the contact rate between two individual household members in population *p*, is the same as observed in Digaale:

$$d_{p0,i} = \frac{d_{00,i}}{s_0} s_p \tag{29}$$

#### Section B. Model fit

Supplemental Table B1 describes the six fitted model parameters. We used the Differential Evolution Markov Chain with snooker updater (DEzs) algorithm implemented in the *BayesianTools* package in *R* to find parameter values that maximized the multinomial log-likelihood comparing observed age-dependent prevalence estimates in the S, VT, NVT, and B compartments with their model-predicted counterparts at steady state.[14]

We ran four independent DEzs chains in parallel, each with 36,000 model iterations distributed over 3 dependent chains, where the first 6,000 iterations were treated as burn-in and discarded. The MCMC acceptance rate was 15.1% and final joint effective sample size 3,054.

**Supplemental Table B1. Fitted model parameters.** Table shows the fitted model parameter, their assumed prior distribution, and the Gelman-Rubin convergence diagnostic.

| Parameter | Description | Prior distribution | Notes | $\hat{R}^1$ (upper CI) |
|---|---|---|---|---|
| beta_1 | Proportion of effective contacts for children aged <5y | $\sim beta(0.1, 10)$ | Vague prior | 1.001 (1.002) |
| beta_2 | Proportion of effective contacts for children aged 5-14y | $\sim beta(0.1, 10)$ | Vague prior | 1.001 (1.001) |
| beta_3 | Proportion of effective contacts for people aged ≥15y | $\sim beta(0.1, 10)$ | Vague prior | 1.001 (1.001) |
| beta_VT_NVT_u5 | Relative transmissibility of VTs compared to NVTs for children <5y | $\sim lognormal(1, 1)$ | Vague prior | 1.001 (1.001) |
| beta_VT_NVT_o5 | Relative transmissibility of VTs compared to NVTs for people ≥5y | $\sim lognormal(1, 1)$ | Vague prior | 1.001 (1.001) |
| competition | Relative reduction in susceptibility to acquiring additional VT/NVT, when carrying NVT/VT | $\sim beta(4.53, 17.14)$ | Informative prior | 1.001 (1.001) |

1. Gelman-Rubin convergence diagnostics, and the upper value of their 95% confidence interval. The multivariate $\hat{R}$ was 1.002.

The posterior model fit to the observed data is shown for 500 posterior samples in Supplemental Figure B1. Our model was able to fit the observed data well with modelled prevalence close to the point estimates of observed prevalence for nearly all data points. The 95% credible intervals of modelled estimates did not include the observed point estimates of the proportion susceptible, carrying VTs, or carrying NVTs for those aged 15-

29y and 6-14y, but remained within the 95% confidence interval around all observed estimates.

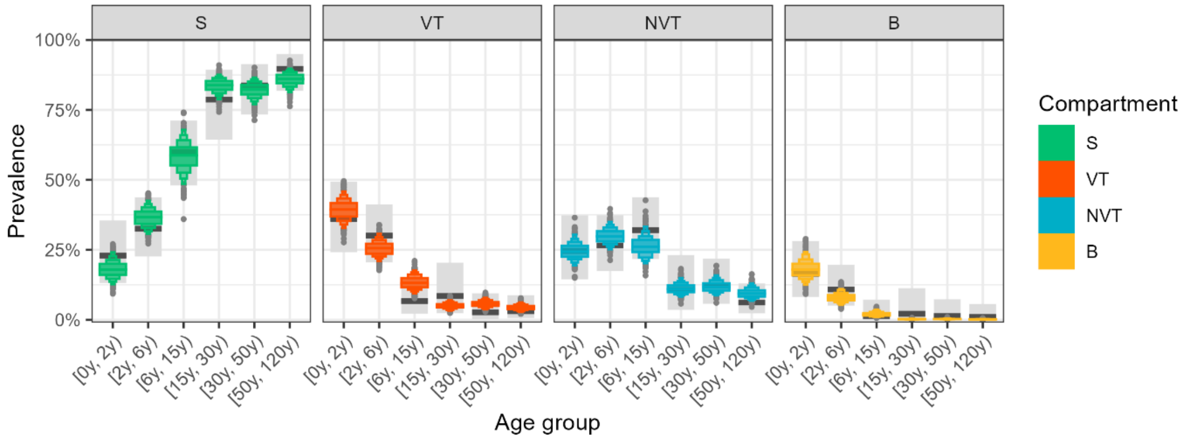

**Supplemental Figure B1. Model fit compared to observed data.** Prevalence by age is shown for S (susceptibles), VT (vaccine-type carriers), NVT (non-vaccine-type carriers) and B (both VT and NVT carriers). Observed estimates are shown as black bars with error bars indicating their 95% confidence intervals. Coloured boxen plots show the distribution of modelled prevalence from 1000 model runs sampling from the joint-posterior distribution.

Thinned trace plots of the four independent chains are shown in Supplemental Figure B2. We achieved good mixing in all chains, that all seemed to have converged. Gelman-Rubin convergence diagnostics ($\hat{R}$) was <1.02 for all fitted parameters (Supplemental Table B1). Posterior and prior marginal density distributions are provided in Supplemental Figure B3. The posterior distribution for the competition parameter is relatively wide which may indicate a reduced identifiability for this parameter value, likely due to the limited amount of data for the prevalence of multiple carriage (B), which was low for all ages ≥6y.

Supplemental Table B2 shows the cross-correlation between the fitted parameters. Values for beta_1 and beta_VT_NVT_u5, that both affect transmission in children <5y, are relatively highly correlated (-0.52). The same is observed for values between the two beta_VT_NVT parameters (-0.83). There is also a relatively strong negative correlation between the competition parameter and beta_1 (-0.62), but not with the other beta parameters, which is probably due to the fact that as for overall prevalence, prevalence of superinfection (B) is highest in the <5y age group.

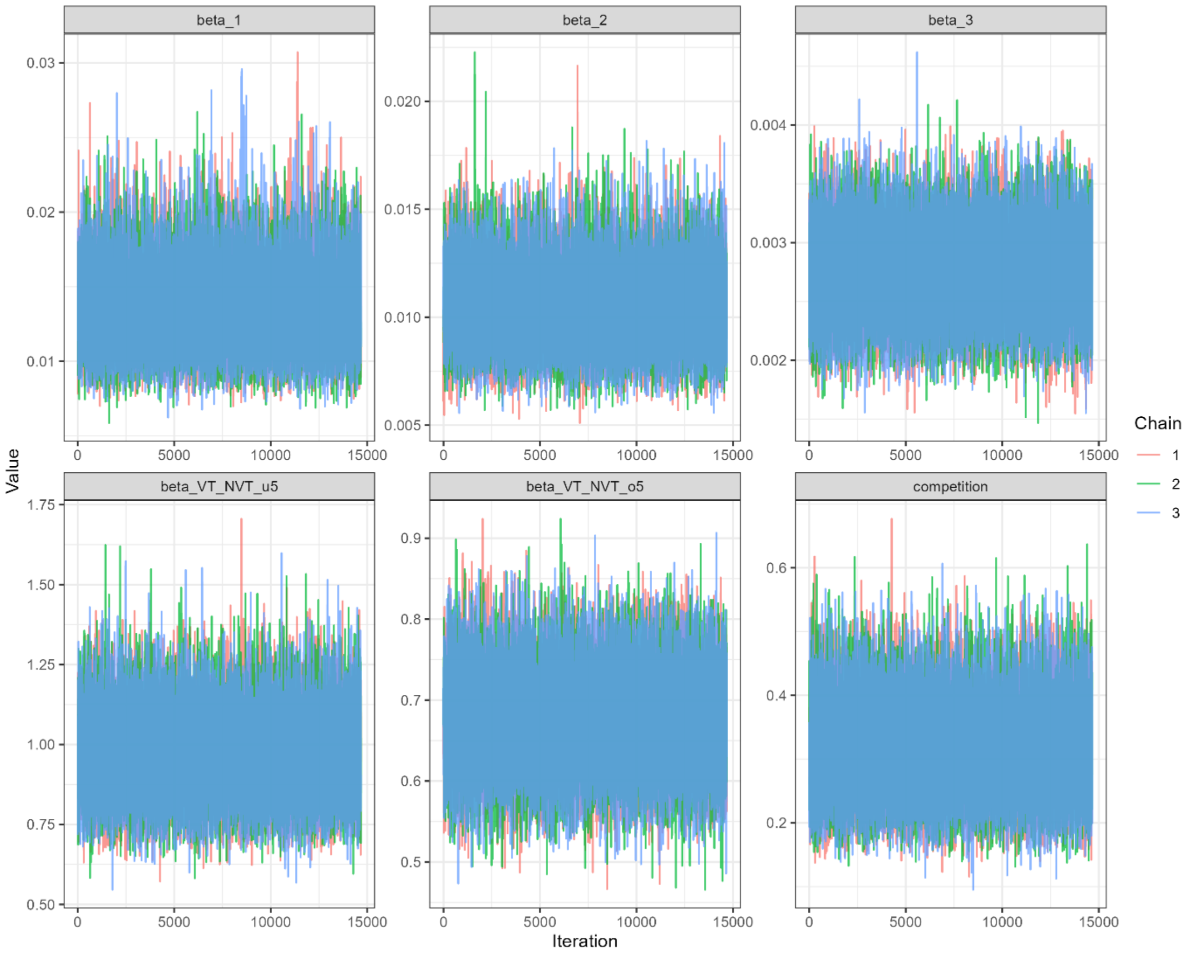

**Supplemental Figure B2. Trace plots of 4 independent chains of fitted model parameters.** Posterior samples are thinned by 10. Posteriors of the dependent DEMC chains within each independent chain are combined.

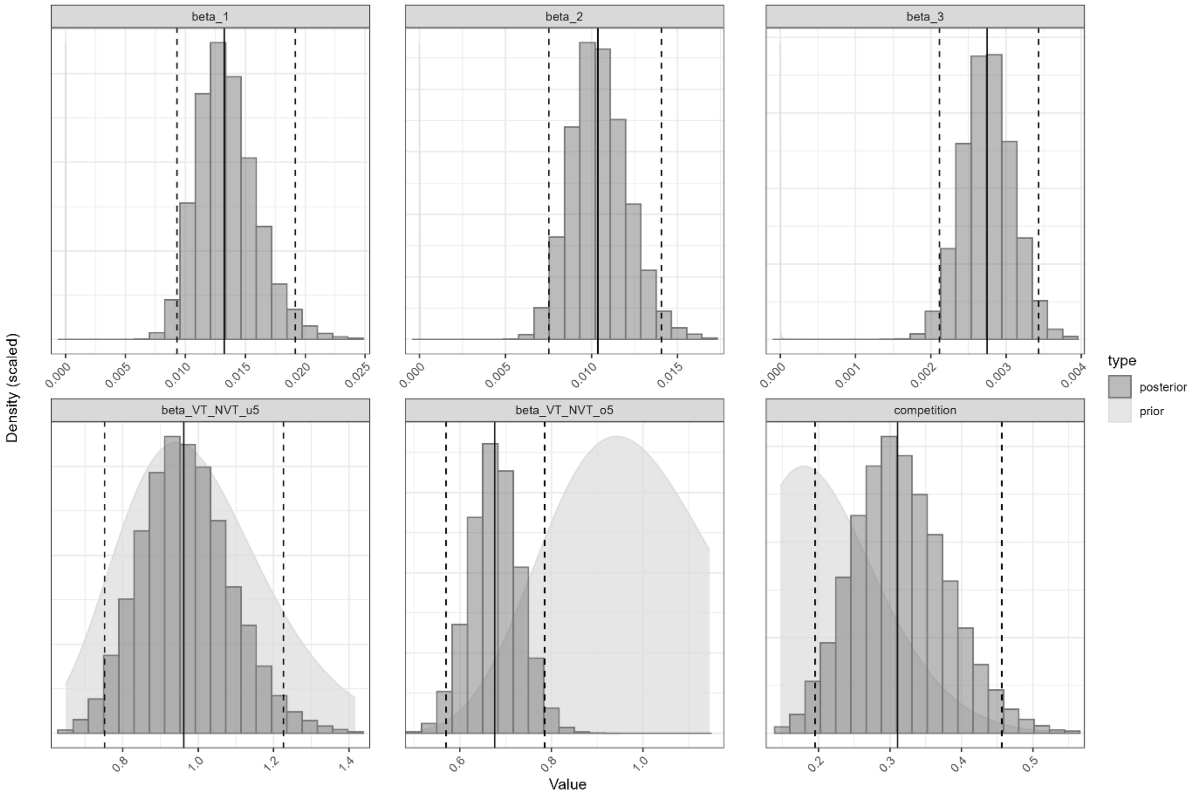

**Supplemental Figure B3. Posterior and prior marginal density distributions of fitted model parameters.** Light- and dark-grey areas show the prior- and posterior density distribution of each parameter. Solid vertical lines indicate the median of the posterior distribution, and dashed vertical lines its 95% credible interval.

**Supplemental Table B2. Cross-correlations of posterior estimates.**

| | beta_1 | beta_2 | beta_3 | beta_VT_NVT_u5 | beta_VT_NVT_o5 | competition |
|---|---|---|---|---|---|---|
| beta_1 | 1 | -0.39 | -0.11 | -0.40 | 0.19 | -0.51 |
| beta_2 | -0.39 | 1 | -0.49 | 0.20 | -0.12 | -0.07 |
| beta_3 | -0.11 | -0.49 | 1 | 0.22 | -0.07 | -0.03 |
| beta_VT_NVT_u5 | -0.40 | 0.20 | 0.22 | 1 | -0.76 | 0.00 |
| beta_VT_NVT_o5 | 0.19 | -0.12 | -0.07 | -0.76 | 1 | -0.08 |
| competition | -0.51 | -0.07 | -0.03 | 0.00 | -0.08 | 1 |

#### Section C. Impact in different settings

*Methods*

While every crisis is unique, crisis-affected populations may be categorized in different typologies: i) mass-displacement in an IDP or refugee camp, typically with high rates of overcrowding and migration, and ii) entrapment in an existing community, which may be rural or urban.[15] To account for a wider diversity of distinct crisis-affected settings, we conducted a sensitivity analysis in which we modelled one exemplar setting for each typology: i) an acute-phase IDP camp based on the Bentiu protection of civilian (PoC) site in South Sudan in 2015, which is much bigger than Digaale, ii) an acute-phase urban setting with mixed IDP and host communities based on the city of Maiduguri in North-East Nigeria in 2016, and iii) a protracted-phase rural setting based on Bambari town, Central African Republic.

Bentiu PoC is a large IDP camp near the town of Bentiu in South Sudan and was established following the start of the 2013-2020 South Sudanese Civil War. When conflict escalated in 2015, the population grew rapidly to 150,000 resulting in overcrowding, poor hygiene, malnutrition.[16]

Maiduguri is the capital of Borno state, Nigeria. Armed conflict in the northeastern states of Nigeria resulted in mass displacement. In 2016, 800,000 displaced individuals were housed with family members, in schools, or in incomplete housing projects throughout the city with a host population of 1 million people. This resulted in high malnutrition rates, high levels of population movement, and high levels of mixing between the displaced and host populations.[17]

Bambari is the capital of the rural Ouaka prefecture in the Central African Republic. It has suffered decades of conflict and instability. While there was no mass displacement and little acute malnutrition in 2019, the protracted crisis has resulted in poor healthcare access with a need to maintain immunity over years with only limited EPI services.[18]

We used data from the 2020 Somaliland Demographic and Health Survey, UN World Population Prospects, and synthetic contact matrices to inform demographic estimates for the host population.[19–21] As we collected detailed data from Digaale, we used it as our base scenario in all analyses.

For each setting, we identified and extracted data on population demographics, malnutrition, and migration (Table 2).[22,23] Our sensitivity analysis then ensures realistic values for these different variables that may occur in these typologies, allowing us to compare the effectiveness of vaccination strategies under different conditions. As in the base model, we modelled a distinct host and displaced population in all settings with the exception of Bambari. In the absence of other pneumococcal prevalence estimates, we assumed prevalence rates in the displaced population to be the same as estimated in Digaale.

**Supplemental Table C1. Scenario specific model parameters.**

| Parameter | Description | Digaale 2019 | Bentiu PoC 2015 | Maiduguri 2016 | Bambari 2019 |
|---|---|---|---|---|---|
| $\sum_{a=1}^{25} N_a^p$ | Total population size in population *p* (combined for malnourished and non-malnourished) | Displaced: 3K [24]; host: 1.2M [19] | Displaced: 150K [25]; host:70K [26] | Displaced: 800K [27]; host: 1M [27] | 375K [22] (no discrepancy made between host and displaced) |
| $h$ | Average household size in displaced population | 4.5 [24] | 7.8 [28] | 5.03 [22] | N/A |
| $c_{a,b}^{p,h}$ | Average number of daily contacts between contactors aged b in population p, and contactees aged a and in location l (within or outside of the household). | Displaced: estimated in Digaale.[24]<br><br>Host: synthetic matrix for Ethiopia.[21] | Displaced: adjusted from Digaale.[24]<br><br>Host: synthetic matrix for South Sudan.[21] | Displaced: adjusted from Digaale.[24]<br><br>Host: synthetic matrix for Nigeria.[21] | Synthetic matrix for the Central African Republic. [21] |
| $T_{qp}$ | Proportion of extra-household contacts made by contactors in the population p with contactees in population q. | 2.7% [24] | Assumed to be the same as in Digaale. | 55.6% (Assumed homogenous mixing). | N/A |
| $M_{qp}$ | Daily migration rate from population p (displaced) to population q (host). | 3.5/10,000/year [24] | 12.6/10,000/year[29] | 13.7/10,000/year[30] | N/A |
| $\chi_p$ | Proportion of population that is acutely malnourished. | Displaced: 27.5% [24]; host: 20.0% [19] | Displaced: 17.9% [29]; host: 17.6% [29] | Displaced: 19.2% [31]; host: 19.2% [31] | 4.2% [32] |

We informed contact rates in the displaced population by extrapolating the contact rates from Digaale, adjusting for the population distribution and household sizes (supplemental material section A). Synthetic contact matrices were used to model contact rates within the host populations.

*Results*

Supplemental Figure C1 shows the effect on VT carriage prevalence in four different settings, while Supplemental Figure C2 shows their daily impact on severe pneumococcal disease cases. Supplemental Table C1 shows the total impact over a 0-1y, 0-2y and 0-3y period on all and infant severe pneumococcal disease cases in the different settings, while Supplemental Table C2 shows the NVN to prevent one severe pneumococcal disease case in the same periods and settings.

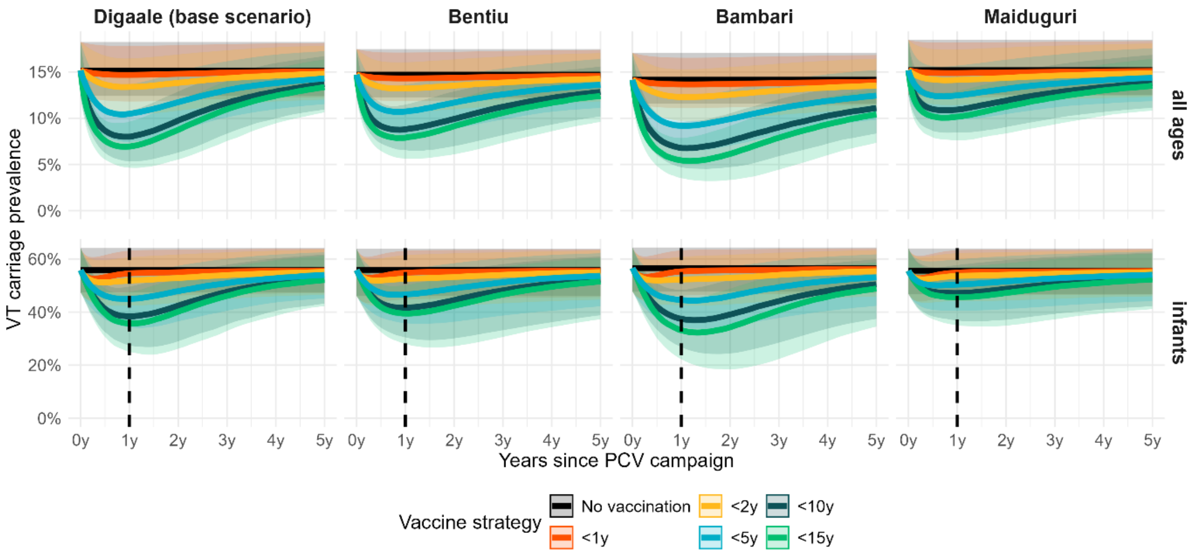

**Supplemental Figure C1. Effect of PCV campaigns on VT carriage prevalence in four settings.** Plots show the effect on VT carriage prevalence over time in infants and all age groups for a single PCV campaign conducted in Digaale, Bentiu, Bambari, and Maiduguri. In all plots, thick lines show the median estimates and shaded areas corresponding 95% credible intervals from 500 model posteriors. A dotted vertical line is plotted for infants 1 year after the PCV campaign. Infants to the right of this line have been born after the PCV campaign and are thus all unvaccinated. The reductions in these birth cohorts result from indirect vaccine protection.

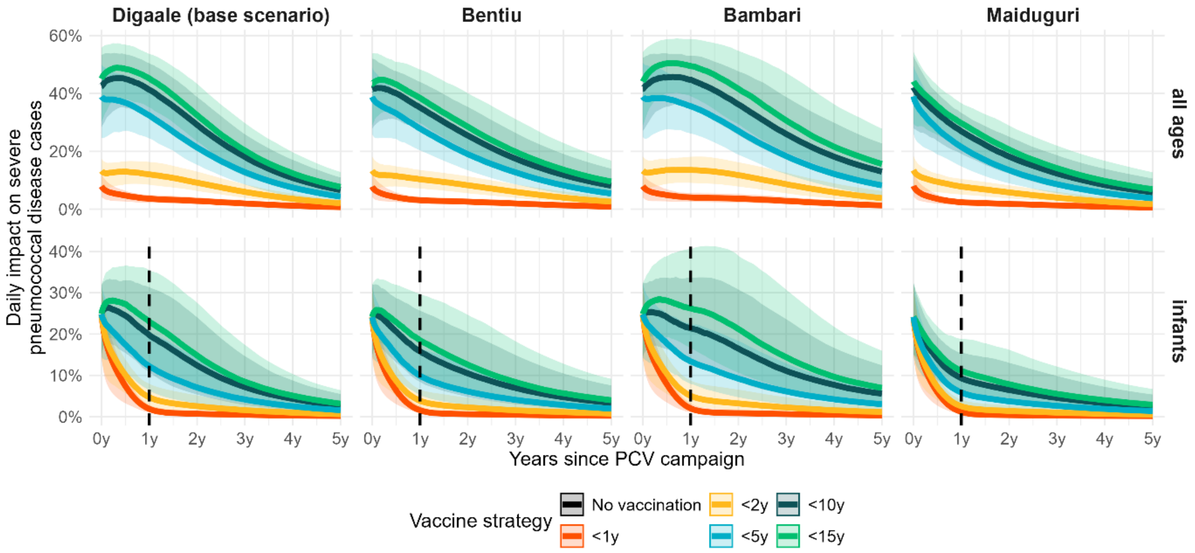

**Supplemental Figure C2. Effect of PCV campaigns on daily severe pneumococcal disease incidence in four settings.** Plots show the daily impact over time in infants and all age groups for campaigns in Digaale, Bentiu, Bambari, and Maiduguri. In all plots, thick lines show the median estimates and shaded areas corresponding 95% credible intervals from 500 model posteriors. A dotted vertical line is plotted for infants 1 year after the PCV campaign. Infants to the right of this line have been born after the PCV campaign and are thus all unvaccinated. The reductions in these birth cohorts result from indirect vaccine protection.

**Supplemental Table C1. Cumulative impact of PCV campaigns on severe pneumococcal disease cases in four different settings.** Estimates show the median absolute cumulative impact per 10,000 people per year, in each respective period. The 95% uncertainty intervals and median relative impact are provided in brackets. Estimates are taken from 500 model runs.

| **Age** | **Vaccine strategy** | **Severe pneumococcal disease cases prevented per 10,000 people per year (median percent reduction)** | | | |
|---|---|---|---|---|---|
| | | *Digaale 2019 (base scenario)* | *Bentiu 2015* | *Maiduguri 2016* | *Bambari 2019* |
| | ***0-1y period*** | | | | |
| *All ages* | <1y | 1 (0 - 1, 5%) | 1 (0 - 1, 4%) | 1 (0 - 1, 4%) | 1 (0 - 1, 5%) |
| | <2y | 2 (1 - 3, 13%) | 2 (1 - 2, 11%) | 1 (1 - 2, 10%) | 2 (1 - 2, 13%) |
| | <5y | 6 (4 - 8, 36%) | 5 (3 - 6, 33%) | 4 (3 - 6, 28%) | 5 (3 - 6, 38%) |
| | <10y | 7 (5 - 9, 44%) | 6 (4 - 8, 40%) | 5 (3 - 7, 34%) | 5 (4 - 8, 45%) |
| | <15y | 7 (5 - 10, 48%) | 6 (4 - 8, 43%) | 5 (4 - 8, 36%) | 6 (4 - 8, 49%) |
| *Infants* | <1y | 13 (6 - 25, 9%) | 11 (5 - 20, 8%) | 10 (5 - 20, 8%) | 11 (5 - 21, 9%) |
| | <2y | 17 (8 - 31, 11%) | 14 (7 - 24, 10%) | 12 (6 - 22, 9%) | 14 (7 - 26, 11%) |
| | <5y | 27 (14 - 50, 18%) | 22 (11 - 37, 16%) | 16 (9 - 30, 12%) | 22 (12 - 41, 18%) |
| | <10y | 37 (20 - 66, 24%) | 29 (15 - 50, 21%) | 21 (11 - 37, 15%) | 29 (16 - 54, 24%) |
| | <15y | 40 (22 - 73, 26%) | 32 (16 - 55, 23%) | 23 (13 - 40, 16%) | 34 (19 - 60, 27%) |
| | ***0-2y period*** | | | | |
| *All ages* | <1y | 1 (1 - 2, 4%) | 1 (1 - 2, 4%) | 1 (1 - 1, 3%) | 1 (1 - 2, 5%) |
| | <2y | 4 (2 - 5, 12%) | 3 (2 - 4, 10%) | 2 (2 - 4, 8%) | 3 (2 - 5, 13%) |
| | <5y | 10 (7 - 14, 32%) | 8 (5 - 11, 28%) | 7 (4 - 10, 23%) | 8 (6 - 12, 35%) |
| | <10y | 12 (9 - 17, 40%) | 10 (7 - 14, 35%) | 8 (6 - 12, 28%) | 10 (7 - 14, 43%) |

| | | | | | |
|---|---|---|---|---|---|
| | <15y | 14 (10 - 19, 43%) | 11 (7 - 15, 38%) | 9 (6 - 13, 30%) | 11 (8 - 16, 48%) |
| *Infants* | <1y | 15 (7 - 28, 5%) | 12 (6 - 22, 4%) | 11 (5 - 21, 4%) | 12 (6 - 24, 5%) |
| | <2y | 22 (11 - 41, 7%) | 18 (9 - 32, 6%) | 15 (7 - 26, 5%) | 19 (10 - 36, 8%) |
| | <5y | 41 (21 - 78, 14%) | 33 (16 - 60, 12%) | 23 (12 - 40, 8%) | 36 (19 - 69, 15%) |
| | <10y | 62 (31 - 115, 20%) | 47 (24 - 87, 17%) | 32 (17 - 57, 11%) | 54 (28 - 102, 21%) |
| | <15y | 70 (36 - 130, 23%) | 54 (27 - 99, 19%) | 36 (19 - 66, 13%) | 65 (34 - 123, 26%) |
| | ***0-3y period*** | | | | |
| *All ages* | <1y | 2 (1 - 2, 4%) | 1 (1 - 2, 3%) | 1 (1 - 2, 3%) | 1 (1 - 2, 4%) |
| | <2y | 5 (3 - 7, 10%) | 4 (3 - 6, 9%) | 3 (2 - 5, 7%) | 4 (3 - 6, 12%) |
| | <5y | 13 (8 - 18, 27%) | 10 (7 - 14, 24%) | 8 (5 - 12, 19%) | 11 (7 - 16, 31%) |
| | <10y | 16 (11 - 23, 34%) | 13 (9 - 18, 30%) | 11 (7 - 15, 24%) | 14 (10 - 20, 39%) |
| | <15y | 18 (13 - 24, 38%) | 14 (10 - 20, 33%) | 12 (8 - 16, 26%) | 16 (11 - 22, 44%) |
| *Infants* | <1y | 16 (8 - 29, 4%) | 13 (6 - 23, 3%) | 12 (6 - 22, 3%) | 13 (6 - 26, 4%) |
| | <2y | 25 (13 - 47, 6%) | 21 (10 - 38, 5%) | 16 (8 - 29, 4%) | 22 (11 - 43, 6%) |
| | <5y | 50 (25 - 96, 11%) | 40 (19 - 74, 10%) | 27 (14 - 47, 7%) | 46 (24 - 90, 12%) |
| | <10y | 76 (37 - 151, 16%) | 59 (29 - 111, 14%) | 39 (21 - 73, 9%) | 71 (36 - 143, 19%) |
| | <15y | 88 (42 - 172, 19%) | 68 (33 - 127, 16%) | 45 (23 - 84, 11%) | 87 (44 - 173, 23%) |

**Supplemental Table C2. Number of vaccines needed to prevent a single severe pneumococcal disease case in four different settings.** Estimates show the total number of PCV doses needed to prevent one case of severe pneumococcal disease. The 95% uncertainty intervals are provided in brackets. Estimates are taken from 500 model runs. Values greater than 1000 are rounded to the nearest hundred.

| Age | Vaccine strategy | NVN: Number of PCV doses needed to prevent one severe pneumococcal disease case | | | |
|---|---|---|---|---|---|
| | | *Digaale 2019 (base scenario)* | *Bentiu 2015* | *Maiduguri 2016* | *Bambari 2019* |
| | ***0-1y period*** | | | | |
| *All ages* | <1y | 327 (209 - 570) | 396 (255 - 736) | 460 (281 - 876) | 389 (246 - 718) |
| | <2y | 265 (193 - 412) | 326 (231 - 517) | 398 (269 - 661) | 318 (221 - 503) |
| | <5y | 238 (173 - 360) | 284 (205 - 440) | 335 (228 - 537) | 281 (197 - 419) |
| | <10y | 406 (299 - 582) | 447 (329 - 663) | 533 (373 - 808) | 463 (331 - 664) |
| | <15y | 549 (396 - 782) | 586 (436 - 869) | 693 (491 - 1,000) | 615 (444 - 882) |
| *Infants* | <1y | 571 (299 - 1,200) | 687 (370 - 1,500) | 722 (376 - 1,600) | 682 (354 - 1,600) |
| | <2y | 958 (517 - 1,900) | 1,100 (650 - 2,200) | 1,300 (710 - 2,700) | 1,100 (609 - 2,300) |
| | <5y | 1,500 (833 - 2,900) | 1,800 (1,100 - 3,500) | 2,400 (1,300 - 4,500) | 1,800 (967 - 3,400) |
| | <10y | 2,300 (1,300 - 4,300) | 2,600 (1,500 - 5,000) | 3,600 (2,000 - 6,500) | 2,700 (1,500 - 4,900) |
| | <15y | 3,100 (1,700 - 5,800) | 3,400 (1,900 - 6,500) | 4,500 (2,600 - 8,200) | 3,400 (1,900 - 6,200) |
| | ***0-2y period*** | | | | |
| *All ages* | <1y | 194 (135 - 328) | 242 (164 - 409) | 302 (194 - 538) | 224 (148 - 372) |
| | <2y | 142 (103 - 219) | 179 (127 - 274) | 233 (159 - 374) | 162 (111 - 246) |
| | <5y | 137 (99 - 205) | 165 (120 - 249) | 209 (142 - 322) | 154 (107 - 223) |
| | <10y | 227 (163 - 324) | 252 (183 - 369) | 321 (228 - 478) | 245 (176 - 343) |
| | <15y | 301 (219 - 425) | 328 (241 - 482) | 409 (296 - 610) | 321 (231 - 453) |
| *Infants* | <1y | 508 (273 - 1,000) | 612 (338 - 1,300) | 672 (354 - 1,400) | 604 (319 - 1,300) |
| | <2y | 723 (391 - 1,400) | 878 (493 - 1,800) | 1,100 (598 - 2,200) | 838 (441 - 1,700) |
| | <5y | 1,000 (532 - 1,900) | 1,200 (667 - 2,500) | 1,700 (968 - 3,300) | 1,100 (576 - 2,100) |
| | <10y | 1,400 (751 - 2,800) | 1,600 (875 - 3,200) | 2,300 (1,300 - 4,400) | 1,500 (771 - 2,800) |
| | <15y | 1,800 (959 - 3,500) | 2,000 (1,100 - 4,000) | 2,900 (1,600 - 5,400) | 1,800 (944 - 3,400) |
| | ***0-3y period*** | | | | |
| *All ages* | <1y | 150 (105 - 249) | 183 (126 - 307) | 238 (158 - 407) | 163 (109 - 267) |
| | <2y | 108 (77 - 166) | 133 (93 - 206) | 179 (123 - 285) | 116 (79 - 177) |
| | <5y | 108 (77 - 162) | 130 (93 - 195) | 169 (115 - 259) | 116 (80 - 170) |
| | <10y | 175 (125 - 250) | 194 (139 - 283) | 251 (181 - 382) | 180 (126 - 255) |
| | <15y | 231 (167 - 326) | 249 (178 - 370) | 318 (225 - 479) | 235 (166 - 333) |
| *Infants* | <1y | 480 (258 - 979) | 576 (323 - 1,200) | 646 (344 - 1,400) | 561 (294 - 1,200) |
| | <2y | 632 (339 - 1,300) | 769 (421 - 1,600) | 976 (541 - 1,900) | 710 (370 - 1,400) |
| | <5y | 827 (434 - 1,700) | 1,000 (540 - 2,100) | 1,400 (827 - 2,700) | 864 (444 - 1,700) |
| | <10y | 1,100 (570 - 2,300) | 1,300 (683 - 2,600) | 1,900 (1,000 - 3,500) | 1,100 (551 - 2,200) |
| | <15y | 1,400 (724 - 3,000) | 1,600 (841 - 3,200) | 2,300 (1,200 - 4,400) | 1,300 (670 - 2,700) |

#### Section D. Impact with routine vaccination coverage

In our base model, we assumed that all populations in our model were PCV-naïve (as is the case in Somaliland, where data was collected). Here, we assessed how estimates change if PCV coverage would have been 20%, 40%, 60%, or 80% rather than 0% prior to the PCV campaign. We assume that the PCV campaign is conducted when routine vaccination breaks down.

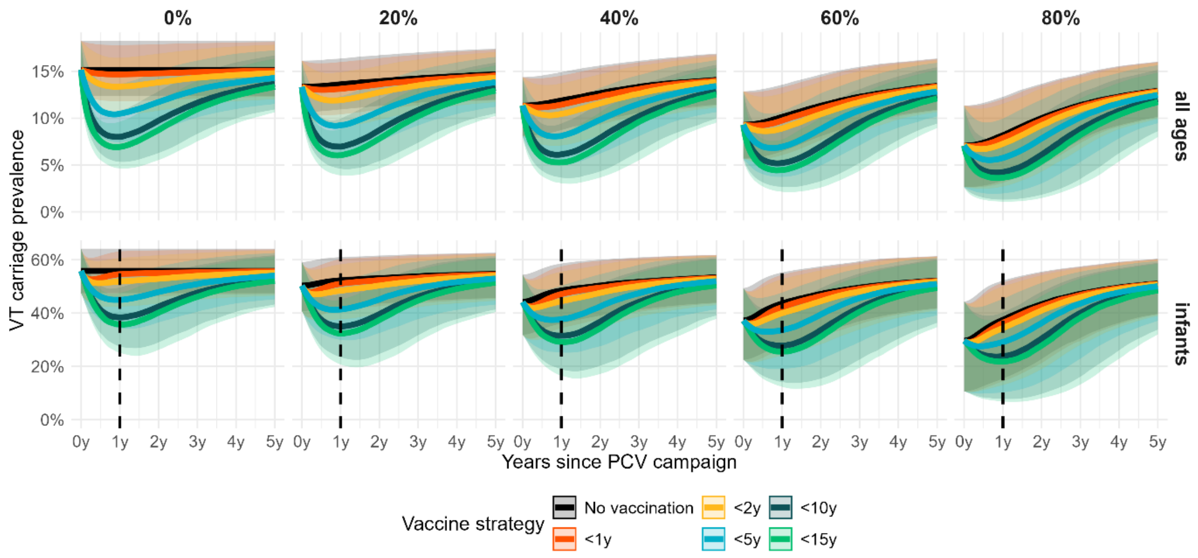

**Supplemental Figure D1. Effect of PCV campaigns on VT carriage prevalence in scenarios with different routine vaccination coverage.** Plots show the effect on VT carriage prevalence over time in infants and all age groups for a single PCV campaign conducted in Digaale in the base scenario of no routine vaccination, and assuming routine PCV coverage at 20-80% pre-PCV campaign. In all plots, thick lines show the median estimates and shaded areas corresponding 95% credible intervals from 500 model posteriors. A dotted vertical line is plotted for infants 1 year after the PCV campaign. Infants to the right of this line have been born after the PCV campaign and are thus all unvaccinated. The reductions in these birth cohorts result from indirect vaccine protection.

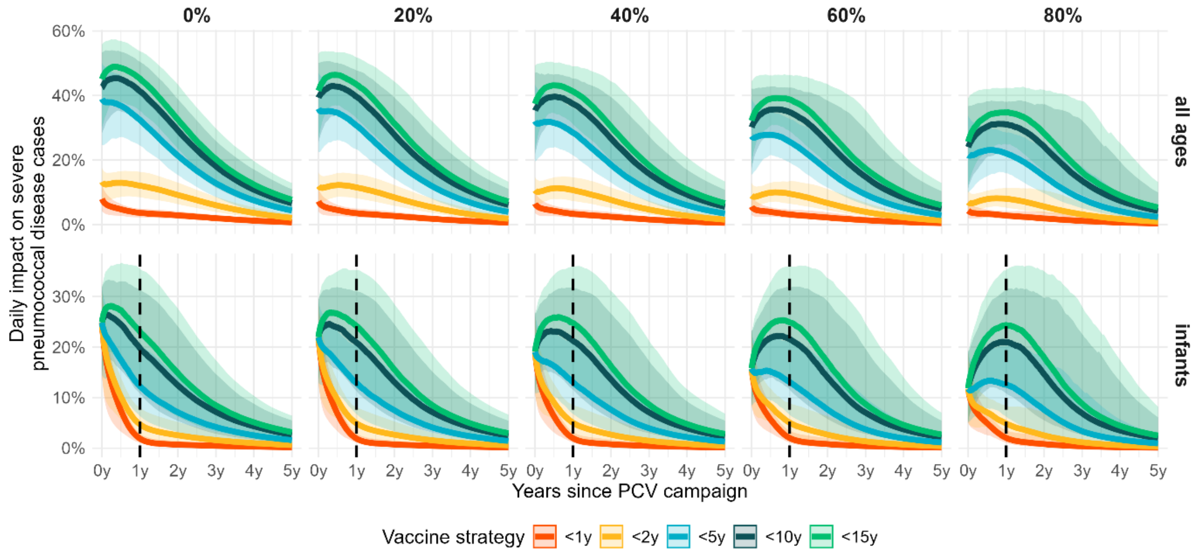

**Supplemental Figure D2. Effect of PCV campaigns on daily severe pneumococcal disease incidence in scenarios with different routine vaccination coverage.** Plots show the daily impact over time in infants and all age groups for a campaign conducted in Digaale in the base scenario of no routine vaccination, and assuming routine PCV coverage at 20-80% pre-PCV campaign. In all plots, thick lines show the median estimates and shaded areas corresponding 95% credible intervals from 500 model posteriors. A dotted vertical line is plotted for infants 1 year after the PCV campaign. Infants to the right of this line have been born after the PCV campaign and are thus all unvaccinated. The reductions in these birth cohorts result from indirect vaccine protection.

**Supplemental Table D1. Cumulative impact of PCV campaigns on severe pneumococcal disease cases in scenarios with different routine vaccination coverage pre-PCV campaign.** Estimates show the median absolute cumulative impact per 10,000 people per year, in each respective period. The 95% uncertainty intervals and median relative impact are provided in brackets. Estimates are taken from 500 model runs.

| **Age** | **Vaccine strategy** | **Severe pneumococcal disease cases prevented per 10,000 people per year** | | | | |
|---|---|---|---|---|---|---|
| | | *0% (base scenario)* | *20%* | *40%* | *60%* | *80%* |
| | ***0-1y period*** | | | | | |
| *All ages* | <1y | 1 (0 - 1, 5%) | 1 (0 - 1, 5%) | 1 (0 - 1, 4%) | 0 (0 - 1, 4%) | 0 (0 - 0, 3%) |
| | <2y | 2 (1 - 3, 13%) | 2 (1 - 2, 12%) | 1 (1 - 2, 11%) | 1 (1 - 1, 10%) | 1 (0 - 1, 8%) |
| | <5y | 6 (4 - 8, 36%) | 5 (3 - 7, 34%) | 4 (3 - 6, 31%) | 3 (2 - 4, 27%) | 2 (1 - 3, 22%) |
| | <10y | 7 (5 - 9, 44%) | 6 (4 - 8, 42%) | 5 (3 - 7, 39%) | 4 (2 - 5, 35%) | 3 (1 - 4, 30%) |
| | <15y | 7 (5 - 10, 48%) | 6 (4 - 9, 45%) | 5 (4 - 7, 42%) | 4 (3 - 6, 38%) | 3 (1 - 5, 33%) |
| *Infants* | <1y | 13 (6 - 25, 9%) | 13 (6 - 25, 9%) | 11 (5 - 21, 8%) | 9 (5 - 18, 7%) | 8 (4 - 14, 6%) |
| | <2y | 17 (8 - 31, 11%) | 17 (8 - 31, 11%) | 15 (7 - 26, 10%) | 13 (7 - 22, 10%) | 10 (5 - 18, 9%) |
| | <5y | 27 (14 - 50, 18%) | 27 (14 - 50, 18%) | 24 (13 - 43, 17%) | 22 (12 - 38, 16%) | 18 (10 - 33, 15%) |
| | <10y | 37 (20 - 66, 24%) | 37 (20 - 66, 24%) | 33 (18 - 58, 23%) | 30 (16 - 53, 22%) | 26 (14 - 47, 21%) |
| | <15y | 40 (22 - 73, 26%) | 40 (22 - 73, 26%) | 37 (20 - 65, 26%) | 33 (18 - 59, 25%) | 29 (15 - 53, 23%) |
| | ***0-2y period*** | | | | | |
| *All ages* | <1y | 1 (1 - 2, 4%) | 1 (1 - 2, 4%) | 1 (1 - 2, 4%) | 1 (1 - 1, 4%) | 1 (0 - 1, 3%) |
| | <2y | 4 (2 - 5, 12%) | 4 (2 - 5, 12%) | 3 (2 - 4, 11%) | 3 (2 - 4, 10%) | 2 (1 - 3, 9%) |
| | <5y | 10 (7 - 14, 32%) | 10 (7 - 14, 32%) | 8 (6 - 12, 30%) | 7 (5 - 10, 27%) | 6 (4 - 8, 24%) |
| | <10y | 12 (9 - 17, 40%) | 12 (9 - 17, 40%) | 11 (8 - 15, 38%) | 9 (7 - 12, 35%) | 7 (5 - 10, 33%) |
| | <15y | 14 (10 - 19, 43%) | 14 (10 - 19, 43%) | 12 (9 - 16, 41%) | 10 (7 - 14, 39%) | 8 (6 - 12, 36%) |
| *Infants* | <1y | 15 (7 - 28, 5%) | 15 (7 - 28, 5%) | 13 (6 - 23, 4%) | 11 (6 - 20, 4%) | 9 (5 - 16, 4%) |
| | <2y | 22 (11 - 41, 7%) | 22 (11 - 41, 7%) | 20 (11 - 35, 7%) | 18 (10 - 31, 6%) | 16 (9 - 27, 6%) |
| | <5y | 41 (21 - 78, 14%) | 41 (21 - 78, 14%) | 39 (19 - 69, 13%) | 36 (18 - 63, 13%) | 32 (16 - 59, 12%) |
| | <10y | 62 (31 - 115, 20%) | 62 (31 - 115, 20%) | 58 (29 - 103, 20%) | 55 (27 - 98, 20%) | 51 (26 - 91, 20%) |
| | <15y | 70 (36 - 130, 23%) | 70 (36 - 130, 23%) | 66 (33 - 116, 23%) | 63 (31 - 111, 23%) | 58 (30 - 103, 23%) |
| | ***0-3y period*** | | | | | |
| *All ages* | <1y | 2 (1 - 2, 4%) | 2 (1 - 2, 4%) | 1 (1 - 2, 3%) | 1 (1 - 2, 3%) | 1 (1 - 1, 3%) |
| | <2y | 5 (3 - 7, 10%) | 5 (3 - 7, 10%) | 4 (3 - 6, 10%) | 3 (2 - 5, 9%) | 3 (2 - 4, 8%) |

| | | | | | | |
|---|---|---|---|---|---|---|
| | <5y | 13 (8 - 18, 27%) | 13 (8 - 18, 27%) | 11 (7 - 15, 25%) | 9 (6 - 12, 23%) | 7 (5 - 10, 20%) |
| | <10y | 16 (11 - 23, 34%) | 16 (11 - 23, 34%) | 14 (10 - 19, 32%) | 12 (9 - 16, 30%) | 10 (7 - 14, 28%) |
| | <15y | 18 (13 - 24, 38%) | 18 (13 - 24, 38%) | 16 (11 - 22, 36%) | 13 (10 - 19, 34%) | 11 (8 - 16, 32%) |
| *Infants* | <1y | 16 (8 - 29, 4%) | 16 (8 - 29, 4%) | 14 (7 - 24, 3%) | 12 (6 - 21, 3%) | 10 (5 - 18, 3%) |
| | <2y | 25 (13 - 47, 6%) | 25 (13 - 47, 6%) | 23 (12 - 41, 5%) | 21 (11 - 37, 5%) | 19 (9 - 32, 5%) |
| | <5y | 50 (25 - 96, 11%) | 50 (25 - 96, 11%) | 47 (22 - 87, 11%) | 44 (21 - 82, 10%) | 40 (19 - 75, 10%) |
| | <10y | 76 (37 - 151, 16%) | 76 (37 - 151, 16%) | 72 (34 - 134, 17%) | 69 (33 - 129, 17%) | 66 (31 - 122, 17%) |
| | <15y | 88 (42 - 172, 19%) | 88 (42 - 172, 19%) | 83 (40 - 154, 19%) | 80 (39 - 150, 19%) | 76 (37 - 141, 19%) |

**Supplemental Table D2. Number of vaccine doses needed to prevent a single severe pneumococcal disease case in scenarios with different routine vaccination coverage pre-PCV campaign.** Estimates show the total number of PCV doses needed to prevent one case of severe pneumococcal disease. The 95% uncertainty intervals are provided in brackets. Estimates are taken from 500 model runs. Values greater than 1000 are rounded to the nearest hundred.

| **Age** | **Vaccine strategy** | **NVN: Number of PCV doses needed to prevent one severe pneumococcal disease case** | | | | |
|---|---|---|---|---|---|---|
| | | *0% (base scenario)* | *20%* | *40%* | *60%* | *80%* |
| | ***0-1y period*** | | | | | |
| *All ages* | <1y | 327 (209 - 570) | 380 (252 - 662) | 461 (302 - 781) | 592 (376 - 972) | 831 (512 - 1,500) |
| | <2y | 265 (193 - 412) | 314 (225 - 493) | 392 (277 - 587) | 516 (360 - 778) | 753 (501 - 1,400) |
| | <5y | 238 (173 - 360) | 287 (197 - 429) | 356 (241 - 541) | 466 (314 - 744) | 655 (411 - 1,500) |
| | <10y | 406 (299 - 582) | 481 (347 - 697) | 587 (421 - 853) | 751 (517 - 1,200) | 1,000 (658 - 2,300) |
| | <15y | 549 (396 - 782) | 641 (463 - 917) | 781 (555 - 1,100) | 991 (686 - 1,500) | 1,300 (853 - 3,000) |
| *Infants* | <1y | 571 (299 - 1,200) | 667 (363 - 1,400) | 801 (428 - 1,600) | 994 (525 - 2,000) | 1,300 (704 - 2,900) |
| | <2y | 958 (517 - 1,900) | 1,100 (624 - 2,200) | 1,300 (721 - 2,400) | 1,500 (875 - 2,900) | 2,000 (1,100 - 4,300) |
| | <5y | 1,500 (833 - 2,900) | 1,700 (970 - 3,100) | 1,900 (1,100 - 3,600) | 2,300 (1,200 - 4,200) | 2,900 (1,500 - 6,300) |
| | <10y | 2,300 (1,300 - 4,300) | 2,600 (1,500 - 4,800) | 2,900 (1,600 - 5,300) | 3,300 (1,800 - 6,300) | 4,100 (2,200 - 9,000) |
| | <15y | 3,100 (1,700 - 5,800) | 3,400 (1,900 - 6,400) | 3,800 (2,100 - 6,900) | 4,300 (2,400 - 8,200) | 5,300 (2,800 - 11,700) |
| | ***0-2y period*** | | | | | |
| *All ages* | <1y | 194 (135 - 328) | 223 (156 - 372) | 266 (187 - 428) | 332 (230 - 501) | 448 (309 - 671) |
| | <2y | 142 (103 - 219) | 169 (120 - 254) | 205 (146 - 301) | 263 (186 - 377) | 363 (257 - 560) |
| | <5y | 137 (99 - 205) | 162 (115 - 232) | 197 (139 - 278) | 247 (174 - 356) | 332 (229 - 588) |
| | <10y | 227 (163 - 324) | 261 (193 - 361) | 309 (226 - 427) | 379 (270 - 537) | 490 (332 - 870) |

| | | | | | | |
|---|---|---|---|---|---|---|
| | <15y | 301 (219 - 425) | 346 (253 - 476) | 407 (296 - 562) | 497 (350 - 702) | 630 (421 - 1,100) |
| *Infants* | <1y | 508 (273 - 1,000) | 585 (330 - 1,200) | 677 (383 - 1,400) | 808 (458 - 1,600) | 1,000 (579 - 2,000) |
| | <2y | 723 (391 - 1,400) | 800 (459 - 1,500) | 886 (513 - 1,700) | 1,000 (586 - 1,900) | 1,300 (715 - 2,400) |
| | <5y | 1,000 (532 - 1,900) | 1,100 (604 - 2,200) | 1,200 (654 - 2,300) | 1,300 (705 - 2,600) | 1,500 (839 - 3,000) |
| | <10y | 1,400 (751 - 2,800) | 1,500 (835 - 3,000) | 1,600 (877 - 3,200) | 1,700 (948 - 3,300) | 1,900 (1,000 - 3,800) |
| | <15y | 1,800 (959 - 3,500) | 1,900 (1,100 - 3,800) | 2,000 (1,100 - 4,000) | 2,200 (1,200 - 4,200) | 2,400 (1,300 - 4,700) |
| | ***0-3y period*** | | | | | |
| *All ages* | <1y | 150 (105 - 249) | 172 (122 - 277) | 205 (146 - 325) | 251 (178 - 388) | 334 (231 - 489) |
| | <2y | 108 (77 - 166) | 127 (90 - 192) | 155 (108 - 228) | 195 (138 - 279) | 267 (188 - 391) |
| | <5y | 108 (77 - 162) | 126 (91 - 184) | 152 (110 - 217) | 190 (135 - 266) | 249 (174 - 372) |
| | <10y | 175 (125 - 250) | 200 (146 - 279) | 234 (171 - 325) | 281 (202 - 395) | 350 (247 - 532) |
| | <15y | 231 (167 - 326) | 262 (190 - 372) | 305 (220 - 429) | 360 (260 - 514) | 444 (307 - 689) |
| *Infants* | <1y | 480 (258 - 979) | 549 (307 - 1,100) | 629 (354 - 1,300) | 739 (422 - 1,400) | 920 (522 - 1,700) |
| | <2y | 632 (339 - 1,300) | 684 (395 - 1,300) | 755 (434 - 1,500) | 856 (498 - 1,700) | 1,000 (599 - 2,100) |
| | <5y | 827 (434 - 1,700) | 877 (479 - 1,900) | 941 (509 - 2,000) | 1,000 (552 - 2,100) | 1,200 (639 - 2,500) |
| | <10y | 1,100 (570 - 2,300) | 1,200 (642 - 2,500) | 1,200 (665 - 2,600) | 1,300 (704 - 2,700) | 1,400 (779 - 3,000) |
| | <15y | 1,400 (724 - 3,000) | 1,500 (808 - 3,100) | 1,600 (831 - 3,200) | 1,600 (884 - 3,400) | 1,800 (985 - 3,700) |

### Section E. Sensitivity analyses

#### *Parametric sensitivity analyses*

We assessed the sensitivity of model outcomes on our assumptions for several key parameters: i) the degree of mixing with the host community, ii) migration rate, iii) relative increased susceptibility of malnourished children, iv) vaccine coverage, v) duration of vaccine protection, and vi) vaccine efficacy against transmission.

As i, ii, and iii affect the pre-vaccination transmission dynamics, we first refitted our model for each new parameter value, running two independent DEzs chains for each model fit, fitting the same parameters using the same priors as provided in Supplemental Table B1. Each independent DEzs chain was ran for 36,000 model iterations distributed over 3 dependent chains, with the first 6,000 iterations were treated as burn-in and discarded. Gelman-Rubin convergence diagnostics ($\hat{\boldsymbol{R}}$) for the fifteen model fits are shown in Supplemental Table E1. $\hat{\boldsymbol{R}}$ values were well < 1.1 or all models.

**Supplemental Table E1. Gelman-Rubin convergence diagnostics of refitted models.** Table shows for each refitted model (rows) the overall and parameter-specific Gelman-Rubin convergence diagnostics (columns).

| Model | Mult. PSRF[a] | β 1 | β 2 | β 3 | β VT/NVT <5y | β VT/NVT ≥5y | Comp |
|---|---|---|---|---|---|---|---|
| *contact_host* | | | | | | | |
| **0** | 1.011 | 1.01 (1.025) | 1.002 (1.005) | 1.004 (1.009) | 1.007 (1.015) | 1.007 (1.017) | 1.006 (1.013) |
| **1.7**[b] | . | . | . | . | . | . | . |
| **5** | 1.012 | 1.011 (1.023) | 1.002 (1.005) | 1.004 (1.01) | 1.004 (1.01) | 1.004 (1.009) | 1.005 (1.011) |
| **10** | 1.017 | 1.003 (1.006) | 1.006 (1.012) | 1.008 (1.02) | 1.003 (1.006) | 1.009 (1.02) | 1.005 (1.012) |
| **20** | 1.014 | 1.007 (1.014) | 1.002 (1.004) | 1.003 (1.007) | 1.003 (1.005) | 1.005 (1.013) | 1.009 (1.02) |
| **40** | 1.014 | 1.007 (1.017) | 1.009 (1.021) | 1.005 (1.013) | 1.007 (1.015) | 1.003 (1.005) | 1.006 (1.014) |
| *malnutrition_transmission* | | | | | | | |
| **1** | 1.015 | 1.007 (1.017) | 1.003 (1.007) | 1.009 (1.018) | 1.004 (1.009) | 1.006 (1.014) | 1.009 (1.022) |
| **1.2**[b] | . | . | . | . | . | . | . |
| **1.4** | 1.007 | 1.006 (1.012) | 1.004 (1.009) | 1.004 (1.01) | 1.005 (1.013) | 1.005 (1.012) | 1.002 (1.003) |
| **1.6** | 1.009 | 1.011 (1.023) | 1.002 (1.004) | 1.003 (1.006) | 1.006 (1.013) | 1.004 (1.01) | 1.006 (1.013) |

| | | | | | | | |
|---|---|---|---|---|---|---|---|
| **1.8** | 1.018 | 1.008 (1.019) | 1.004 (1.009) | 1.003 (1.007) | 1.012 (1.023) | 1.008 (1.018) | 1.008 (1.018) |
| **2** | 1.012 | 1.012 (1.026) | 1.007 (1.018) | 1.003 (1.007) | 1.004 (1.008) | 1.005 (1.012) | 1.008 (1.02) |
| *Migration* | | | | | | | |
| **0** | 1.015 | 1.005 (1.012) | 1.002 (1.004) | 1.006 (1.016) | 1.008 (1.018) | 1.008 (1.019) | 1.003 (1.006) |
| **2** | 1.012 | 1.007 (1.014) | 1.003 (1.007) | 1.002 (1.006) | 1.006 (1.014) | 1.004 (1.009) | 1.001 (1.004) |
| **4** | 1.01 | 1.008 (1.017) | 1.006 (1.014) | 1.008 (1.02) | 1.01 (1.021) | 1.008 (1.019) | 1.006 (1.014) |
| **6** | 1.014 | 1.006 (1.014) | 1.004 (1.009) | 1.009 (1.021) | 1.002 (1.005) | 1.005 (1.009) | 1.009 (1.019) |
| **7.7[b]** | . | . | . | . | . | . | . |
| **10** | 1.01 | 1.01 (1.02) | 1.007 (1.013) | 1.006 (1.014) | 1.009 (1.02) | 1.007 (1.017) | 1.003 (1.007) |

Gelman-Rubin convergence diagnostics, and the upper value of their 95% confidence interval, for the six fitted parameters: beta_1, beta_2, beta_3, beta_VT_NVT_u5, beta_VT_NVT_o5, competition.

a. The upper value of the multivariate $\widehat{\boldsymbol{R}}$ is not shown.
b. The main model as not refitted

For each fitted model (including the baseline model), we simulated 250 post-vaccination runs sampling from their joint posterior and compared the univariate impact of the changed parameter value on six summary statistics: the maximum impact on infant VT prevalence, number of days since PCV campaign at which peak impact on infant VT prevalence is reached, the cumulative 3y impact on severe pneumococcal disease cases in infants and all ages, and their NVN, for each vaccine strategy <1y, <2y, <5y, <10y, and <15y, shown in Supplemental Figures E1 and E2.

*Mixing with host population*

Increasing the proportion of extra-household contacts made with the host community reduced the overall impact of vaccination on all outcomes. In general, it makes the impact of the different strategies more similar, reducing the additional benefit of wider age targeting with increased mixing with the host population, presumably as indirect protection is limited. The most drastic change is observed on the maximum impact of prevalence, and the number of days at which impact on prevalence peaks. Values for the <5y and <15y strategies shift from a peak impact of 17% (11 – 22) after 306 (191 – 441) days and 34% (22

– 47) after 366 (242 – 472) days in the absence of mixing with the host population, to 10% (8 – 11) after 149 (127 – 177) days and 15% (12 – 19) after 181 (152 – 1206) days when 40% of extra-household contacts are made with members of the host population. The reduced impact is also observed on the reduced impact on cumulative severe pneumococcal disease cases: the impact of <5y and <15y campaigns on all cases during the 3y following the campaign reduces from preventing 28% (25 – 32) and 42% (35 – 51) in the absence of mixing with the host population, to 22% (19 – 25) and 32% (28 – 35). While extended age targeting remains beneficial with increased mixing with the host population, the impact on infant cases is smaller. This difference reflects the fact that i) a substantial disease burden remains in older age groups, and that ii) carriers in older ages contribute to the transmission to those older age groups, in contrast to infants. While campaigns become less efficient when mixing with the host population increases, the change in efficiency is minimal.

*Migration rate*

Migration removes vaccinated individuals from the population and introduces new unvaccinated individuals. Therefore, increasing the migration rate reduces the overall impact of the different vaccination strategies. It reduces the relative difference between the strategies, though the overall impact of wider age targeting remains substantially higher for all migration values considered.

In the absence of migration, peak impact on infant VT carriage prevalence occurs 426 (304 – 544) days following the PCV campaign with a strategy targeting children <15y, and 368 (233 – 514) for an <5y campaign. These values reduce to 358 (233 – 483) and 296 (184 – 452) days when individuals remain displaced for, on average, 10 years, while they reduce even further to 238 (179 – 296) and 200 (147 – 261) days if individuals would move in, on average, 2 years. Migration also affects the maximum impact on infant VT prevalence. An

<15y campaign could reduce VT prevalence by 36% (25 – 49) in the absence of migration but would only reduce it by 24% (16 – 32) if migration occurs after 2 years (on average).

In the absence of migration, an <5y campaign would be able to prevent up to 32% (28 – 36) of all severe pneumococcal disease cases over a 3-year period. If individuals would migrate after, on average, 2 years, an <15y campaign would be required to achieve the same effect (28%, 23 – 32). In the absence of migration, the same <15y campaign would prevent 48% (41 – 53) of all cases over the 3-year period.

Migration especially affects the longevity of the impact of PCV campaigns with wider age targeting. Over a 2-year period, the relative difference between the scenarios with the highest considered migration rate and the absence of migration was 32% for an <15y campaign (a 36% compared to a 53% reduction). However, the cumulative impact of an <15y campaign over 5-years would be 45% (38 – 52) in the absence of migration, but only 21% (18 – 23) with the highest migration rate, a relative difference of 53%.

*Increased acquisition malnourished*

Increasing the relative acquisition rate of malnourished compared to non-malnourished individuals did not substantially affect any of the modelled outcomes. There may be a very small reduction in impact as the relative acquisition rate increases, most obvious in the cumulative 5y impact in infants, but it is a negligible effect.

This is not very surprising, as we refitted our models to the same carriage prevalence estimates in all models. We assume that carriage prevalence is an average of the total population (malnourished and non-malnourished) and refitting the model with the same proportion of the population that is malnourished, but an increased acquisition rate, has adjusted the fitted beta parameters so that the modelled overall prevalence remains close to the observed estimates. We would expect an increase in overall carriage prevalence if the acquisition rate of a segment of the population would increase, but we don't have reliable data to parameterize such models accurately.

*Vaccine coverage*

Vaccine coverage affects both direct and indirect vaccine protection. Lower vaccine coverage would result in reduced vaccine impact, although the duration until peak impact of infant VT prevalence would be achieved is not substantially affected, especially for vaccine strategies with narrower age targeting. A vaccine coverage of only 45% would result in a peak impact prevalence after 252 (188 – 371) days and 287 (216 – 385) for an <5 and <15y campaign, whilst this would occur after 309 (224 – 423) and 360 (267 – 453) days if vaccine coverage would be 95%. In contrast, it would substantially affect the maximum impact on infant VT prevalence that would be achieved, from 20% (15 – 28) and 37% (25 – 48) for <5y and <15y campaigns with 95% coverage, to only 9% (6% - 12%) and 16% (11 – 22) for the same campaigns with 45% coverage.

Again, scenarios that result in lower indirect effects, in this case lower vaccine coverage, would reduce the impact of all strategies, but would especially diminish the additional benefit of increased age targeting. An <5y campaign with 95% coverage would prevent 31% (28 – 36) of all severe pneumococcal disease cases over a 3y period, but only 15% (13 – 18) at 45% coverage. In contrast, an <15y campaign at 95% coverage would prevent 43% (37 – 50) of all severe pneumococcal disease cases, but only 22% (19 – 26) at 45% coverage.

Similarly, impact on severe pneumococcal disease cases would only be 6% (4 – 8) and 9% (6 - 13) for an <5y and <15y campaign at 45% coverage, down from 13% (10 – 18) and 21% (14 – 29) at 95% coverage. This result it is not surprising but highlights the need for high vaccine coverage to maximize impact.

*Duration of vaccine protection*

The duration of vaccine protection primarily affects the longevity of the PCV campaign. While we have good estimates for the duration of vaccine protection of PCV, these are based on 3 or 4 dose schedules in infants, given in routine schedules. It is possible that the average duration of vaccine protection is lower following a reduced dose schedule, suboptimal vaccine storage, or in vaccinees with certain health conditions such as malnutrition. Our results do not suggest that the impact of a PCV campaign is very sensitive for this assumption, except for very low values for the duration of vaccine protection, that are probably unrealistic. As for migration rates, the assumed duration of vaccine protection primarily affects the longevity of a PCV campaign, e.g. the impact over a 3y period or longer.

*Vaccine efficacy against transmission*

Finally, we assessed the sensitivity of our results against the assumed VE against transmission. Again, we have good estimates for the VE of PCVs, but it may be lower following a reduced dose schedule -especially in very young children, suboptimal storage or delivery of vaccine, or in people with certain underlying conditions. We also assessed a scenario where we assumed VE_c to be 0, not because we think this is realistic, but to highlight the importance of the indirect effect of the PCV campaign.

Unsurprisingly, reduced VE against transmission would result in substantially reduced impact of a PCV campaign. This by itself highlights the importance of the indirect effects on pneumococcal transmission. If PCVs would not protect against transmission, there would be no added benefit of widening the age targeting of a campaign on infant severe pneumococcal disease cases. However, as not all cases occur in infants, an <5y campaign would still prevent 20% (16 – 23) of cases over a 3y period, whereas an <10y and <15y campaign would increase this to 24% (20 – 27) and 25% (21 – 28). This highlights the importance of direct protection, even for these older age groups.

However, the assumed VE against carriage of 50% increases the impact of an <5y campaign on all severe pneumococcal disease cases over 3y to 28% (25 – 32) and increases the impact of an <15y campaign to 39% (34 – 46). The indirect protection from the wider age targeting even increases the impact on infant cases over 3y from 4% (3 – 4) for both vaccine strategies to 11% (9 – 16) and 18% (13 – 26) for the <5y and <15y campaigns.

To summarise, our sensitivity analyses show that our modelled results of the impact of PCV campaigns are especially sensitive to factors that affect the indirect effect of the PCV campaign: the assumed contact rates with the (unvaccinated) host population, vaccine coverage, and vaccine efficacy against transmission. Wider age targeting could mitigate the reduced impact from any of these factors, but this may come at a cost of reduced efficiency. The impact is also affected by the migration rate and the duration of vaccine protection, but to lower degree than the other factors. These especially affect the longevity of the impact of the PCV campaign, beyond a 3y period. Finally, our results are not affected by the assumed increased acquisition rate in malnourished individuals, primarily because we lack the data to accurately parameterize this variable.

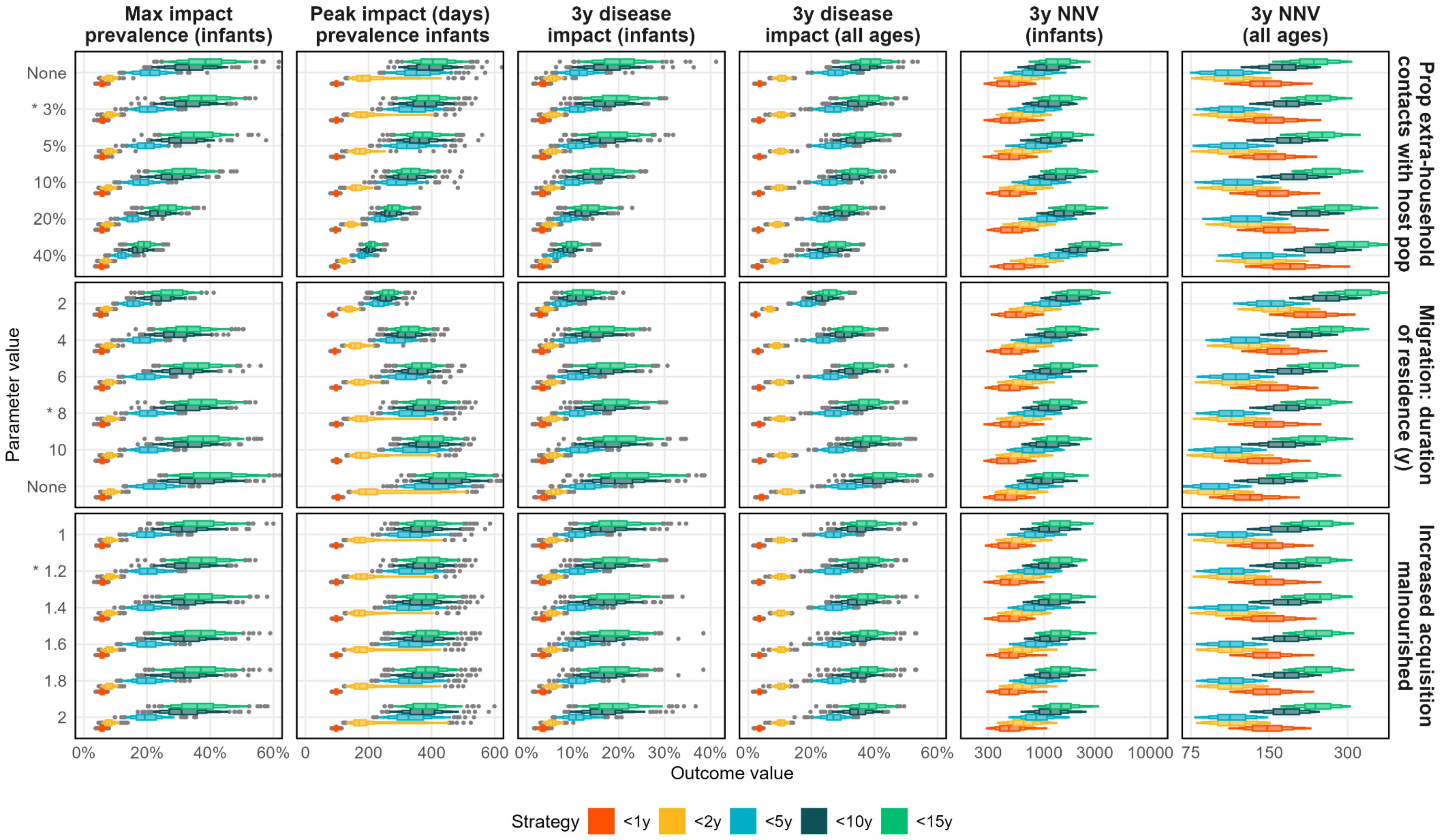

**Supplemental Figure E1. Parametric sensitivity analyses for the mixing with the host population, migration rate, and relative susceptibility of malnourished individuals.** Parameters are shown in facet rows, with parameter values on the y-axes. Parameter values denote i) the proportion of contacts made outside of the crisis-affected population, ii) the average number of years individuals remain resident in the crisis-affected population, and iii) the relative increase in susceptibility to acquisition of pneumococci in malnourished individuals. Parameter values used in the main model (1.7%, 7.7 years, and 1.2) are highlighted with an asterisk. Different outcomes are shown in facet columns, with outcome values on the x-axes. Outcome values denote i) the maximum relative reduction on infant VT prevalence, ii) the time (days) after the PCV campaign at which i occurs, iii) the cumulative impact on severe pneumococcal disease over three years following the PCV campaign in all age groups and infants, and iv) the NVN for these campaigns. Boxen plots show the distribution of the outcome values (x-axes) of 100 model runs for vaccine strategies targeting children <1y (red), <2y (yellow), <5y (blue), <10y (dark green), and <15y (light green).

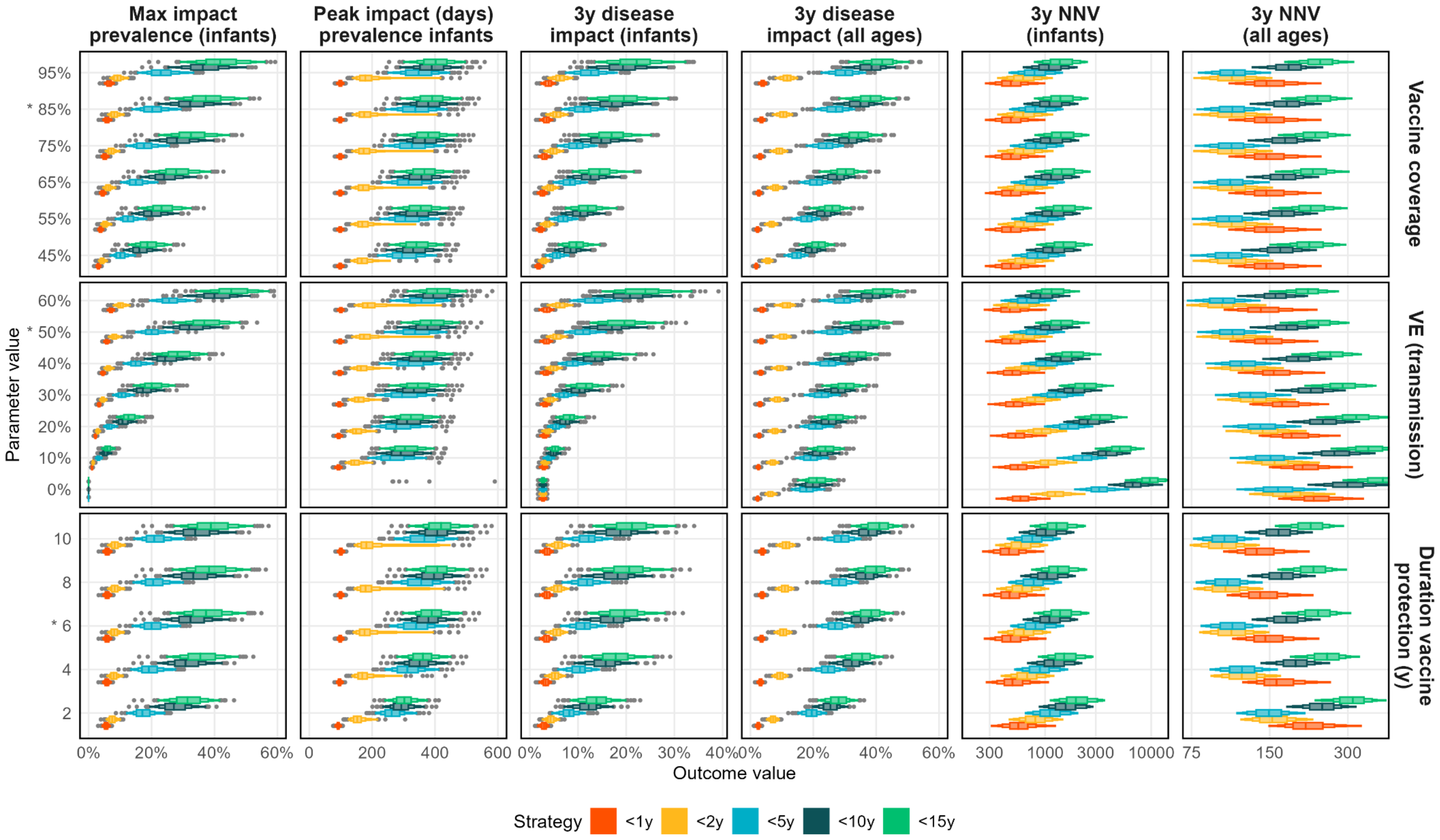

**Supplemental Figure E2. Parametric sensitivity analyses for the vaccine coverage, duration of vaccine protection, and vaccine efficacy.** Parameters are shown in facet rows, with parameter values on the y-axes. Parameter values denote i) the vaccine coverage in targeted age groups, ii) the average duration of vaccine protection in years, and the vaccine efficacy against transmission. Parameter values used in the main model (85%, 8 years, and 50%) are highlighted with an asterisk. Different outcomes are shown in facet columns, with outcome values on the x-axes. Outcome values denote i) the maximum relative reduction on infant VT prevalence, ii) the time (days) after the PCV campaign at which i occurs, iii) the cumulative impact on severe pneumococcal disease over three years following the PCV campaign in all age groups and infants, and iv) the NVN for these campaigns. Boxen plots show the distribution of the outcome values (x-axes) of 100 model runs for vaccine strategies targeting children <1y (red), <2y (yellow), <5y (blue), <10y (dark green), and <15y (light green).

*Alternative model structure*

Our main model structure assumes no superinfections of serotypes of the same group (VT or NVT), and only promotes coexistence of serotypes arbitrarily. We explored the structural uncertainty of our model to this assumption by assessing our results in an alternative structurally neutral model that does allow for superinfection of serotypes of the same group (Supplemental Figure E3).

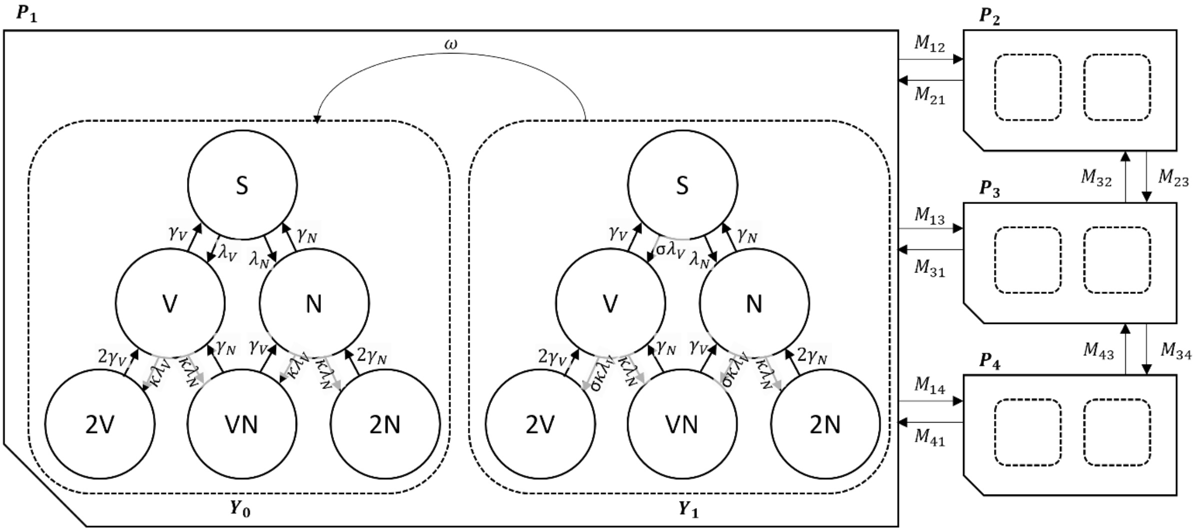

**Supplemental Figure E3. Compartmental model structure of the pneumococcal transmission model.** Pneumococcal serotypes defined as those included in the PCV vaccine (vaccine-type, V), or not included in the vaccine (non-vaccine type, N). At any time $t$, Susceptible individuals (S) acquire serotypes in either group at rates $\lambda_V(t)$ and $\lambda_N(t)$, and immediately become infectious carriers themselves. Carriers of VTs or NVTs remain partially susceptible to infection with an additional serotype, and may develop a superinfection (2V, VN, 2N) at rates $\kappa\lambda_V(t)$ and $\kappa\lambda_N(t)$, where $\kappa$ represents competition between serotypes. Carriers are assumed to clear their infection of all serotypes in either group at rates $\gamma_V$ for VTs and $\gamma_N$ for NVTs. Individuals may be without vaccine derived protection ($Y_0$), or with full vaccine derived protection ($Y_1$). Those with vaccine protection acquire VT at a reduced rate $\sigma$, and lose their level of protection at rate $\omega$. Different populations $P_1$, $P_2$, and $P_3$, $P_4$ represent malnourished and non-malnourished strata of a displaced and host population. Individuals of population $p$ migrate to population $q$ at rate $M_{qp}$. Age groups, ageing, and transmission between vaccine protection strata, populations, and age groups are not shown.

*Epidemiological transitions*

In this alternative model structure, the values for epidemiological compartment $X_a^{p,y}$, where $X \in \{S, V, N, 2V, VN, 2V\}$ (i.e. $S_a^{p,y}$, $V_a^{p,y}$, $N_a^{p,y}$, $2V_a^{p,y}, VN_a^{p,y}, 2N_a^{p,y}$), again hold the proportion of people in age group *a* in population *p*, that are in compartment *X* in vaccinated stratum *y*.

Values for model compartments within a single age group in a model population sum to 1, i.e.

$$\sum_{y=0}^{N_{\text{vac}}} \left( S_a^{p,y} + V_a^{p,y} + N_a^{p,y} + 2V_a^{p,y} + VN_a^{p,y} + 2N_a^{p,y} \right) = 1 \qquad 30$$

The demographic changes in the model compartments, $\Delta D_{X_a^{p,y}}(t)$, remain the same as in the base model structure listed in equation 4. The epidemiological transitions are altered as shown in the following set of equations. For ease of reading, we have again omitted the time notations, and gave the notations for the age group, population, and vaccinated stratum a lighter colour:

$$\Delta E_{S_a^{p,y}}(t) = -\left(\sigma_a^{p,y} \lambda_{V_a}^{\ p} + \lambda_{N_a}^{\ p}\right)\overline{S}_a^{p,y} + \gamma_{V_a}\overline{V}_a^{p,y} + \gamma_{N_a}\overline{N}_a^{p,y}$$

$$\Delta E_{V_a^{p,y}}(t) = \sigma_a^{p,y} \lambda_{V_a}^{\ p}\overline{S}_a^{p,y} - \left(\sigma_a^{p,y} \kappa\lambda_{V_a}^{\ p} + \kappa\lambda_{N_a}^{\ p} + \gamma_{V_a}\right)\overline{V}_a^{p,y} + \gamma_{N_a}\overline{VN}_a^{p,y} + 2\gamma_{V_a}\overline{2V}_a^{p,y}$$

$$\Delta E_{N_a^{p,y}}(t) = \lambda_{N_a}^{\ p}\overline{S}_a^{p,y} - \left(\kappa\lambda_{N_a}^{\ p} + \sigma_a^{p,y} \kappa\lambda_{V_a}^{\ p} + \gamma_{N_a}\right)\overline{N}_a^{p,y} + \gamma_{V_a}\overline{VN}_a^{p,y} + 2\gamma_{N_a}\overline{2N}_a^{p,y}$$

$$\Delta E_{2V_a^{p,y}}(t) = \sigma_a^{p,y} \kappa\lambda_{V_a}^{\ p}\overline{V}_a^{p,y} - 2\gamma_{V_a}\overline{2V}_a^{p,y}$$

$$\Delta E_{VN_a^{p,y}}(t) = \kappa\lambda_{N_a}^{\ p}\overline{V}_a^{p,y} + \sigma_a^{p,y} \kappa\lambda_{V_a}^{\ p}\overline{N}_a^{p,y} - \left(\gamma_{V_a} + \gamma_{N_a}\right)\overline{VN}_a^{p,y}$$

$$\Delta E_{2N_a^{p,y}}(t) = \kappa\lambda_{N_a}^{\ p}\overline{N}_a^{p,y} - 2\gamma_{N_a}\overline{2N}_a^{p,y}$$

$$\lambda_{V_a}^{\ p} = \beta_a^{p,V} \sum_{b=1}^{N_{\text{age}}} \sum_{q=1}^{N_{\text{pop}}} \left( c_{ba}^{p}\left(\delta_{q,p}(1 - o^p) + o^p T_{qp}\right) \sum_{y=0}^{N_{\text{vac}}} \left( \overline{V}_b^{q,y} + \overline{2V}_b^{q,y} + \overline{VN}_b^{q,y} \right) \right)$$

$$\lambda_{N_a}^{\ p} = \beta_a^{p,N} \sum_{b=1}^{N_{\text{age}}} \sum_{q=1}^{N_{\text{pop}}} \left( c_{ba}^{p}\left(\delta_{q,p}(1 - o^p) + o^p T_{qp}\right) \sum_{y=0}^{N_{\text{vac}}} \left( \overline{N}_b^{q,y} + \overline{2N}_b^{q,y} + \overline{VN}_b^{q,y} \right) \right) \qquad 31$$

The parameter definitions and values are the same as in the main model. We then combine the demographic and epidemiological transitions, and calculate the change for each model compartment at time *t* as:

$$\frac{dX_a^{p,y}}{dt} = \Delta D_{X_a^{p,y}}(t) + \Delta E_{X_a^{p,y}}(t) \qquad 32$$

*Model fit*

Supplemental Figure E4 shows the fit of the alternative model compared to the observed data. We got an overall good fit to the data, with the exception of prevalence in <2y in the VT2 compartment, which was overestimated in the model.

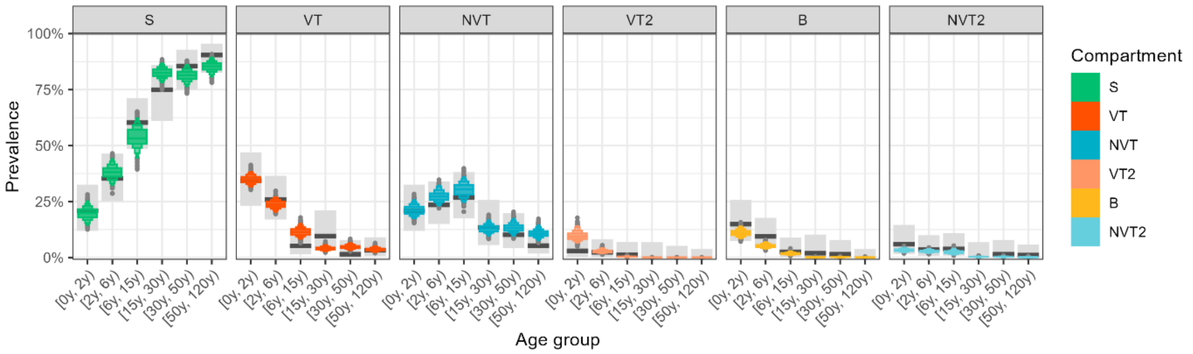

**Supplemental Figure E4. Model fit to observed data.** Prevalence by age is shown for S (susceptibles), VT (vaccine-type carriers), NVT (non-vaccine-type carriers), VT2 (carriers of 2 or more VTs, but no NVTs), B (both VT and NVT carriers), and NVT2 (carriers of 2 or more NVTs, but no VTs). Observed estimates are shown as black bars with error bars indicating their 95% confidence intervals. Coloured boxen plots show the distribution of modelled prevalence from 1000 model runs sampling from the joint-posterior distribution.

*Model outcomes*

Supplemental Figure E5 shows the impact of different PCV campaigns on VT prevalence and severe pneumococcal disease incidence over time. Supplemental Figure E6 shows the NVN to prevent one severe pneumococcal disease case for different age groups and periods since the PCV campaign.

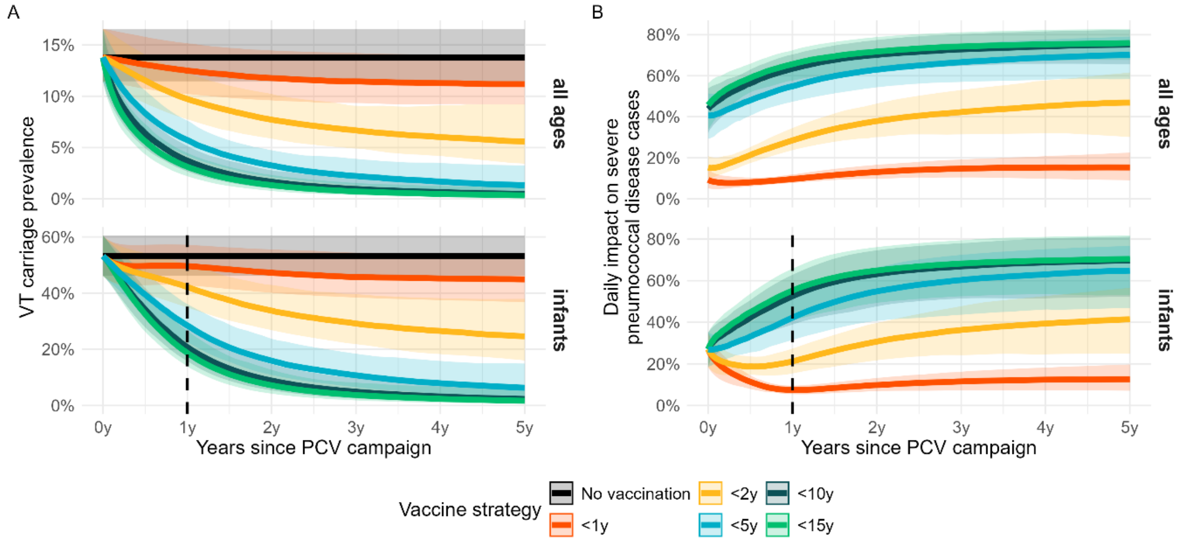

**Supplemental Figure E5. Impact of PCV mass vaccination campaigns on prevalence and incidence.** A: VT carriage prevalence (VT + 2VT + B) in all age groups and infants only. B: reduction in daily severe pneumococcal disease cases compared to no vaccination, in all age groups and infants only. In all plots, thick lines show the median estimates and shaded areas corresponding 95% credible intervals from 500 model posteriors. A dotted vertical line is plotted for infants 1 year after the PCV campaign. Infants to the right of this line have been born after the PCV campaign and are thus all unvaccinated. The reductions in these birth cohorts result from indirect vaccine protection.

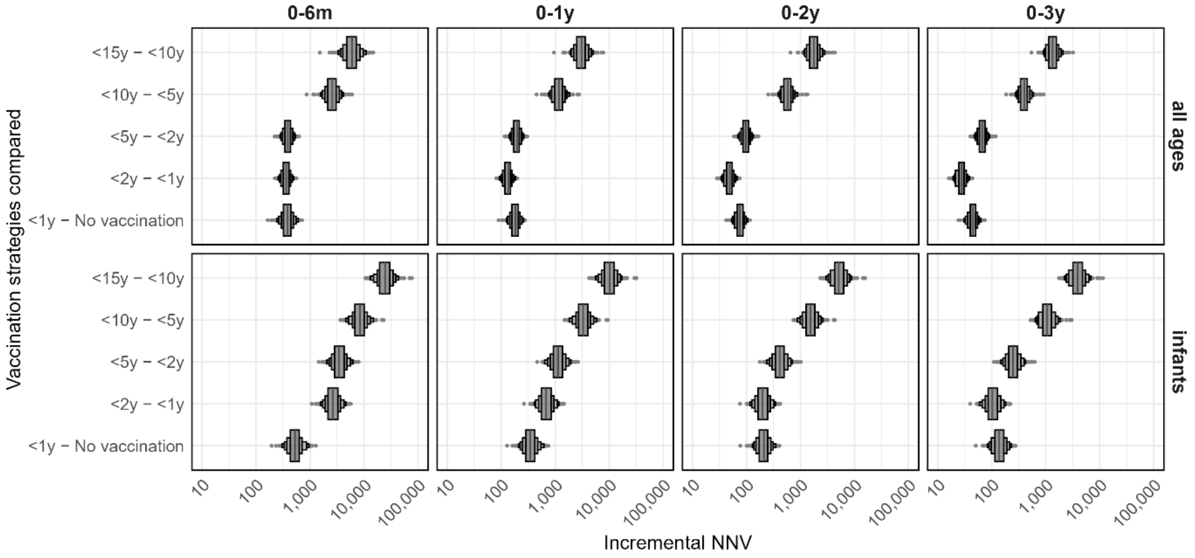

**Supplemental Figure E6. Number needed to vaccinate to prevent one severe pneumococcal disease case.** In different time periods (facet columns) and age groups (facet rows). Estimates show median and 95% CrI from 500 posterior samples.

Our alternative model projected overall much greater impact of all PCV campaigns, with much longer duration. However, qualitatively, this did not alter the relative rank of each vaccination strategy in terms of impact and efficiency compared to our base model. The base model structure has been used to effectively explain pneumococcal transmission dynamics before and after PCV introduction in several settings[1,11,33,34], while the neutral model has predominantly been used to understand coexistence of biologically similar strains.[35,36] We therefore retained the former as the main model in our analysis.

#### Section F. Global demand forecast

*Rationale*

In order to more fully illuminate the case for PCV use by humanitarian actors, and to understand the unmet need, we used our identified vaccination strategies to estimate the potential demand for PCV over the next five years.

*Methods*

We extracted estimates of the number of displaced individuals from the last five years from the child displacement dataset from Unicef, which aggregates data from UNHCR refugee statistics, the Global Internal Displacement Database, and UNRWA.[37–40] We assumed that it would be unlikely that PCV campaigns would be needed in high income settings, where refugees would likely be covered by routine immunization schedules, and restricted the dataset to countries classified as low or lower-middle income countries by the World Bank.[41] As the Unicef displacement dataset does not provide age-specific data, we took population estimates for the included countries from UN WPP, assumed the age distributions were representative for the displaced population, and used these values to estimate the age-specific proportions of the displaced populations, e.g. the total number of children <5y.[20]

We then assumed that all children <5y or <10y in currently displaced populations would receive a single dose of PCV. We assumed that the average increasing trend in displaced populations would continue, and that each newly displaced population would receive an <5y or <10y PCV campaign as well. We assumed a vaccination coverage of 100% in our calculations, without any adjustment for wasting of vaccine doses. We ignored potential maintenance strategies after the PCV campaign.

*Results*

In 2024, an estimated 87 million people were forcibly displaced in low and lower middle-income countries, which has on average increased by 16 million people annually within the past five years. On average, 14% and 27% of these populations are aged <5y and <10y.

28 million doses of PCV would be needed if an <5y campaign would be implemented in all these populations. If <10y campaigns would be implemented, 55 million doses of PCV would be needed. If two doses would be administered in infants, the need would increase to 34 and 60 million doses.

While these are very rough estimates assuming PCVs would be used in all displaced populations, they highlight the substantial need there would be in order to make PCVs available to crisis-affected populations.

#### References used in supplemental material

1 Flasche S, Ojal J, Le Polain de Waroux O, *et al.* Assessing the efficiency of catch-up campaigns for the introduction of pneumococcal conjugate vaccine: a modelling study based on data from PCV10 introduction in Kilifi, Kenya. *BMC Med* 2017; **15**: 113.

2 Awori JO, PERCH Study Group. Association between malnutrition and pneumococcal pneumonia in the PERCH study. 2018. https://publichealth.jhu.edu/sites/default/files/2024-02/9isppd-511ax.pdf.

3 von Mollendorf C, Cohen C, de Gouveia L, *et al.* Risk Factors for Invasive Pneumococcal Disease Among Children Less Than 5 Years of Age in a High HIV Prevalence Setting, South Africa, 2010 to 2012. *The Pediatric Infectious Disease Journal* 2015; **34**: 27.

4 von Mollendorf C, von Gottberg A, Tempia S, *et al.* Increased Risk for and Mortality From Invasive Pneumococcal Disease in HIV-Exposed but Uninfected Infants Aged <1 Year in South Africa, 2009–2013. *Clinical Infectious Diseases* 2015; **60**: 1346–56.

5 Verhagen LM, Hermsen M, Rivera-Olivero IA, *et al.* Nasopharyngeal carriage of respiratory pathogens in Warao Amerindians: significant relationship with stunting. *Tropical Medicine & International Health* 2017; **22**: 407–14.

6 Verhagen LM, Gómez-Castellano K, Snelders E, *et al.* Respiratory infections in Eñepa Amerindians are related to malnutrition and Streptococcus pneumoniae carriage. *J Infect* 2013; **67**: 273–81.

7 Gebre T, Tadesse M, Aragaw D, *et al.* Nasopharyngeal Carriage and Antimicrobial Susceptibility Patterns of Streptococcus pneumoniae among Children under Five in Southwest Ethiopia. *Children (Basel)* 2017; **4**: 27.

8 County Department of Health. Kilifi County SMART Survey Report. 2016. https://www.nutritionhealth.or.ke/wp-content/uploads/SMART%20Survey%20Reports/Kilifi%20County%20SMART%20Survey%20Report%20November2016.pdf.

9 Hammitt LL, Etyang AO, Morpeth SC, *et al.* Effect of ten-valent pneumococcal conjugate vaccine on invasive pneumococcal disease and nasopharyngeal carriage in Kenya: a longitudinal surveillance study. *The Lancet* 2019; **393**: 2146–54.

10 Silaba M, Ooko M, Bottomley C, *et al.* Effect of 10-valent pneumococcal conjugate vaccine on the incidence of radiologically-confirmed pneumonia and clinically-defined pneumonia in Kenyan children: an interrupted time-series analysis. *The Lancet Global Health* 2019; **7**: e337–46.

11 Ojal J, Griffiths U, Hammitt LL, *et al.* Sustaining pneumococcal vaccination after transitioning from Gavi support: a modelling and cost-effectiveness study in Kenya. *The Lancet Global Health* 2019; **7**: e644–54.

12 Lipsitch M, Abdullahi O, D'Amour A, *et al.* Estimating rates of carriage acquisition and clearance and competitive ability for pneumococcal serotypes in Kenya with a Markov transition model. *Epidemiology* 2012; **23**: 510–9.

13 Melegaro A, Choi YH, George R, Edmunds WJ, Miller E, Gay NJ. Dynamic models of pneumococcal carriage and the impact of the Heptavalent Pneumococcal Conjugate Vaccine on invasive pneumococcal disease. *BMC Infectious Diseases* 2010; **10**: 90.

14 Ter Braak CJF, Vrugt JA. Differential Evolution Markov Chain with snooker updater and fewer chains. *Stat Comput* 2008; **18**: 435–46.

15 Checchi F, Warsame A, Treacy-Wong V, Polonsky J, Ommeren M van, Prudhon C. Public health information in crisis-affected populations: a review of methods and their use for advocacy and action. *The Lancet* 2017; **390**: 2297–313.

16 UN OCHA. South Sudan | OCHA. 2023; published online Dec 11. https://www.unocha.org/south-sudan (accessed Dec 18, 2023).

17 UN OCHA. Nigeria | OCHA. 2023; published online Nov 21. https://www.unocha.org/nigeria (accessed Dec 18, 2023).

18 Center for Preventative Action. Conflict in the Central African Republic. Global Conflict Tracker. https://cfr.org/global-conflict-tracker/conflict/violence-central-african-republic (accessed Dec 18, 2023).

19 Central Statistics Department, Ministry of Planning and National Development, Somaliland Government, Ministry of Planning and National Development, Somaliland Government. The Somaliland Health and Demographic Survey 2020. 2020.

20 United Nations, Department of Economic and Social Affairs, Population Division. World Population Prospects - Population Division - United Nations. 2019. https://population.un.org/wpp/Download/Standard/Population/ (accessed April 19, 2020).

21 Prem K, Zandvoort K van, Klepac P, *et al.* Projecting contact matrices in 177 geographical regions: An update and comparison with empirical data for the COVID-19 era. *PLOS Computational Biology* 2021; **17**: e1009098.

22 IOM. Displacement Tracking Matrix. https://dtm.iom.int/ (accessed Dec 18, 2023).

23 UN OCHA. Situation Reports. https://reports.unocha.org/ (accessed Dec 18, 2023).

24 van Zandvoort K, Bobe MO, Hassan AI, *et al.* Social contacts and other risk factors for respiratory infections among internally displaced people in Somaliland. *Epidemics* 2022; **41**: 100625.

25 South Sudan: Cumulative Registration Data, January 2014 - August 2015 - South Sudan | ReliefWeb. 2015; published online Aug 31. https://reliefweb.int/report/south-sudan/south-sudan-cumulative-registration-data-january-2014-august-2015 (accessed April 16, 2024).

26 Checchi F, Testa A, Warsame A, Bs LQ, Burns R. Estimates of crisis-attributable mortality in South Sudan, December 2013-April 2018. 2018.

27 UN OCHA. Northeast Nigeria: Humanitarian emergency - Situation Report No. 1. 2016; published online Nov 28. https://www.unocha.org/publications/report/nigeria/northeast-nigeria-humanitarian-emergency-situation-report-no-1-28-november-2016 (accessed April 16, 2024).

28 South Sudan National Bureau of Statistics (NBS). National Baseline Household Survey 2009 Report. 2012.

29 Concern. Bentiu PoC - Nutrition Anthropometry & Retrospective Mortality Survey Report. 2015. https://info.undp.org/docs/pdc/Documents/SSD/20160124-

180405_CWW%20-%20Bentiu%20PoC%20-%20Smart%20Survey%20-%20August%202015.pdf (accessed April 16, 2024).

30 Action Against Hunger. Report of Small-Scale SMART Survey in MMC, Jere LGAs, Borno. 2016.

31 Nigeria National Bureau of Statistics. Nutrition and food security surveillance: north east Nigeria - Emergency survey. 2019. https://fscluster.org/sites/default/files/documents/nfss_round_8_final_report_november_2019.pdf (accessed April 16, 2024).

32 Unicef. Multiple Indicator Cluster Surveys. https://mics.unicef.org/surveys (accessed April 16, 2024).

33 Le Polain De Waroux O, Edmunds WJ, Takahashi K, *et al.* Predicting the impact of pneumococcal conjugate vaccine programme options in Vietnam. *Hum Vaccin Immunother* 2018; **14**: 1939–47.

34 Choi YH, Jit M, Flasche S, Gay N, Miller E. Mathematical Modelling Long-Term Effects of Replacing Prevnar7 with Prevnar13 on Invasive Pneumococcal Diseases in England and Wales. *PLOS ONE* 2012; **7**: e39927.

35 Gjini E, Valente C, Sá-Leão R, Gomes MGM. How direct competition shapes coexistence and vaccine effects in multi-strain pathogen systems. *J Theor Biol* 2016; **388**: 50–60.

36 Lipsitch M, Colijn C, Cohen T, Hanage WP, Fraser C. No coexistence for free: Neutral null models for multistrain pathogens. *Epidemics* 2009; **1**: 2–13.

37 United Nations High Commissioner for Refugees. Global Trends. UNHCR Global Trends. https://www.unhcr.org/global-trends (accessed May 16, 2025).

38 Global Internal Displacement Database. IDMC - Internal Displacement Monitoring Centre. https://www.internal-displacement.org/database (accessed Dec 19, 2023).

39 UNRWA Registered Population Dashboard. UNRWA. https://www.unrwa.org/what-we-do/relief-and-social-services/unrwa-registered-population-dashboard (accessed Dec 19, 2023).

40 Child Displacement and Refugees. UNICEF DATA. https://data.unicef.org/topic/child-migration-and-displacement/displacement/ (accessed Dec 19, 2023).

41 World Bank Country and Lending Groups – World Bank Data Help Desk. https://datahelpdesk.worldbank.org/knowledgebase/articles/906519-world-bank-country-and-lending-groups (accessed Dec 19, 2023).
